# Lack of dietary fibre increases gut microbiome-derived uremic toxins that contribute to increased blood pressure

**DOI:** 10.1101/2025.09.16.25335853

**Authors:** Chudan Xu, Liang Xie, Christopher K. Barlow, Leticia Camargo Tavares, Evany Dinakis, Panayiotis Louca, Julia El-Sayed Moustafa, Chaoran Yang, Michael Nakai, Xiaosuo Wang, Giovanni Guglielmi, Dakota Rhys-Jones, Joanne A. O’Donnell, Stephanie Yiallourou, Melinda J. Carrington, Gavin W. Lambert, Jane Muir, Charles R. Mackay, Darren J. Creek, David M. Kaye, Kerrin Small, John O’Sullivan, Cristina Menni, Francine Z. Marques

## Abstract

Insufficient dietary fibre intake is a known risk factor for high blood pressure and cardiovascular disease, yet its mechanistic basis remains unclear. Here, we identify a gut microbial pathway linking fibre deprivation to elevated blood pressure. In mice, low-fibre diets shifted microbial resource preference toward tyrosine fermentation, increasing host exposure to p-Cresol-derived metabolites, particularly p-Cresol glucuronide (PCG). Oral L-tyrosine, the precursor for p-Cresol, modestly increased PCG under normal fibre conditions, while antibiotics abolished it. In two healthy human cohorts, lower fibre intake was associated with higher PCG, which correlated with elevated blood pressure and co-expression with immune pathways. Mendelian Randomisation analysis supported a causal relationship between PCG and blood pressure. In a randomised controlled trial, fibre supplementation reduced both circulating PCG and blood pressure in individuals untreated for hypertension. These findings reveal a microbiota-mediated mechanism by which fibre deficiency promotes tyrosine fermentation and PCG production, contributing to elevated blood pressure.

## Introduction

Hypertension affects approximately 1.3 billion people globally and remains the leading modifiable risk factor for death and cardiovascular disease (CVD).^1,2^ It is also a major contributor to chronic kidney disease (CKD), accounting for 43.2% of age-standardised CKD disability-adjusted life years (DALYs) in 2017.^3^ Persistent high blood pressure (BP) damages renal microvasculature, impairing excretory function and promoting uraemic solute retention.^4^ Moreover, hypertension and CKD are bidirectionally linked: declining renal function elevates BP, perpetuating a vicious cycle.^5^

Lifestyle interventions, particularly dietary changes, are first-line strategies for managing hypertension.^6^ Dietary fibre reduces BP and lowers CVD and CKD risk and mortality.^7–9^ A now well-accepted phenomenon involves microbial fermentation of fibre into short-chain fatty acids (SCFAs), such as acetate and butyrate,^10–14^ which modulate host physiology through G protein-coupled receptor activation and anti-inflammatory pathways.^15–18^ However, global fibre intake remains suboptimal—averaging ∼11g/day versus recommended levels of 25–30g/day—posing risks not only for hypertension but also for gut microbiota dysbiosis, as many types of fibre are prebiotic and essential in maintaining a healthy microbiota.^19–21^

Despite growing evidence linking fibre, SCFAs, and BP regulation, the consequences of fibre deficiency on microbial metabolite production and host BP remain poorly defined. We hypothesised that low fibre intake promotes the generation of harmful microbial metabolites that contribute to hypertension. Using both experimental and clinical samples, we show that fibre deficiency shifts gut microbial metabolism toward tyrosine fermentation, increasing production of *p*-Cresol-derived compounds—particularly *p*-Cresol glucuronide (PCG). We demonstrate that PCG is causally associated with elevated BP in humans, and that an SCFA-enriched fibre randomised clinical trial that reduced BP also lowers PCG levels. These findings uncover a previously unrecognised microbial pathway linking fibre deficiency to hypertension and suggest that targeting microbial *p*-Cresol production may offer a novel therapeutic strategy for BP control, with broad implications for cardiovascular and renal health.

## Results

### Dietary fibre modulated the mouse gut microbiome and plasma metabolome

To investigate the impact of dietary fibre on the gut microbiome and the host metabolome in hypertension, we fed mice a low- or high-fibre diet under sham (normal BP) or Ang II (high BP) conditions (Fig.1A). We previously reported that low-fibre mice developed higher BP and cardiac dysfunction than high-fibre-fed Ang II-mice, and that this effect was transferable using faecal microbiota transplantation to germ-free mice.^12^ Using 16S rRNA amplicon sequencing, we found dietary fibre, and not Ang II, was the most significant modulator of the gut microbiome (Supplementary Fig.1). We observed similar results using shotgun sequencing, showing fibre was the key driver of changes in taxonomic abundances (Supplementary Fig.2A-B). In particular, a low-fibre diet enriched the microbiome for *Anaerotruncus* and *Oscillibacter*, both bacterial genera from the Oscillospiraceae family. In contrast, a high-fibre diet enriched the microbiome for *Bacteroides intestinalis* and *Clostridium symbiosum* (Supplementary Fig.2B). We identified 299 and 1,007 gene families (Supplementary data: Fig.2C and Table 1), and 22 and 55 enzymes (Enzyme Commission, EC) were differentially enriched in the caecal content from low- and high-fibre groups, respectively (Supplementary data: Fig.2D and Table 2).

**Figure 1.**
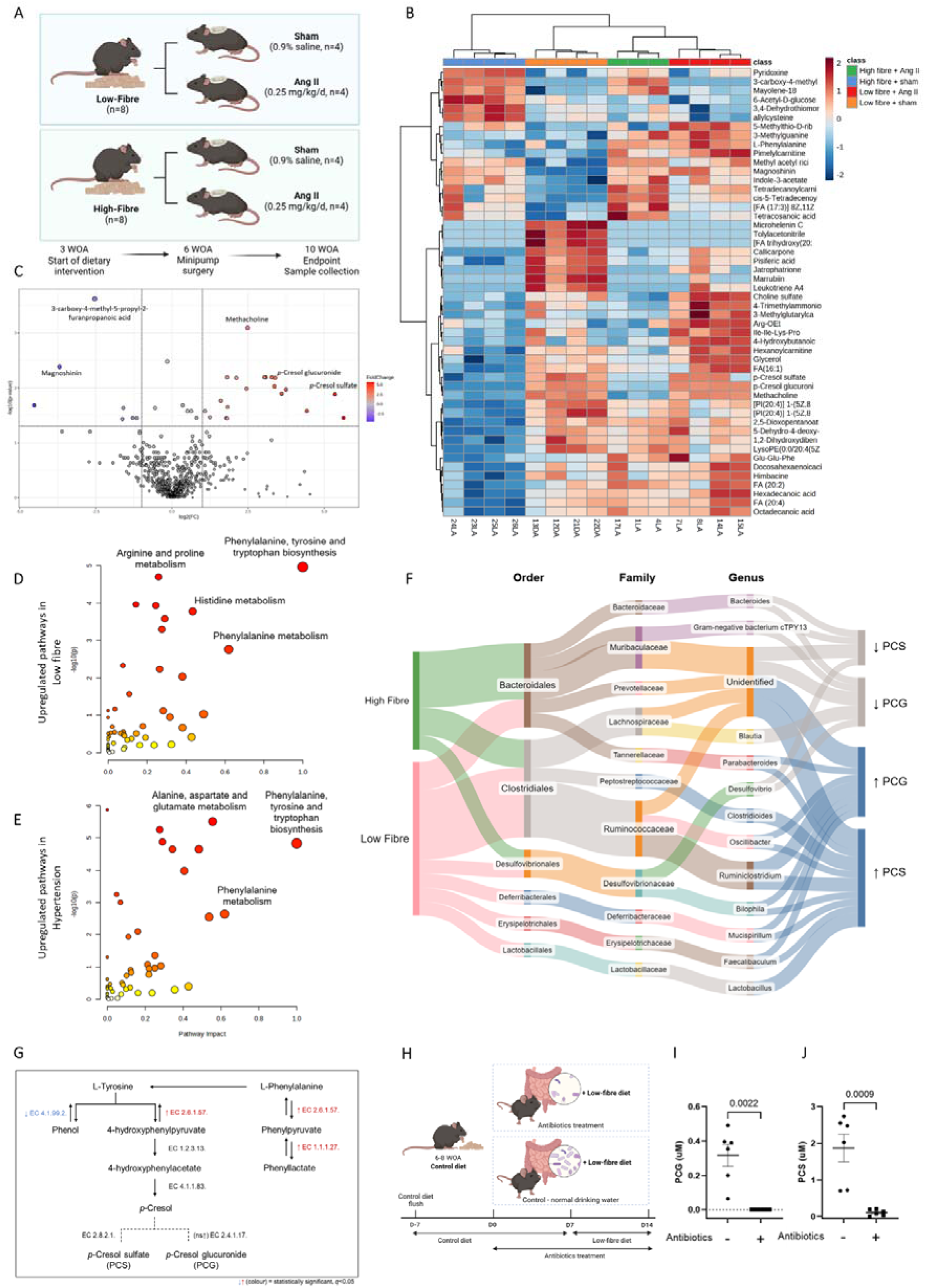
Insufficient dietary fibre increased gut-derived uremic toxins and altered host metabolomic pathways. **(A)** Experimental design. Mice had either low- or high-fibre dietary intervention at three-week-old of age (WOA). At 6 WOA, they had minipump surgery and received either Ang II intervention (hypertensive group) or saline (sham/normotensive group) (n=3-4/group). After 4 weeks of intervention, plasma was collected and used for metabolomic profiling. **(B)** Heatmap showing the top 50 putatively annotated metabolites relevant among groups (see methods). **(C)** Volcano plot showing statistically different metabolites between low- and high-fibre intake in sham mice. Upregulated pathways in **(D)** low-fibre compared to high-fibre-fed mice (irrespective of the surgery intervention), and **(E)** hypertensive compared to normotensive mice (irrespective of dietary intervention) (n=7-8/group). **(F)** The bacterial taxa associated with low- and high-fibre intake and the levels of *p*-Cresol glucuronide (PCG) and *p*-Cresol sulfate (PCS) in the host metabolome. **(G)** Changes in microbial enzyme genes involved in L-Tyrosine metabolism from mice on a low-fibre diet. Only enzymes of interest are shown here; enzyme genes related to these metabolisms are not limited to the details provided (EC, enzyme commission – KEGG database). Full arrows: metabolisms through bacterial fermentation; dotted arrows: metabolisms through enzymatic reactions of the host. **(H)** Antibiotics study design to determine whether metabolites of interest are dependent on the gut microbiota. Mice were randomised and placed under either normal drinking water or water treated with an antibiotic cocktail of enrofloxacin (10mg/kg body weight/day) and amoxicillin with clavulanic acid (50mg/kg body weight/day) to reduce the gut bacterial load until the end of the experiment (n=6 per group). The relative abundance of **(I)** PCG and **(J)** PCS (uM) in the plasma was measured by targeted LC-MS (n=6/group). Experimental design of Ang II-induced hypertensive mice on dietary fibre intervention. Legends: Ang II, angiotensin II; LC-MS, liquid chromatography-mass spectrometry. **(B-E)** Analyses were performed via MetaboAnalyst using one-way ANOVA, FDR<0.05 and |log2FC|≥1. The pathway enrichment analysis was based on KEGG Pathway. Ang II, angiotensin II. KEGG, Kyoto Encyclopedia of Genes and Genomes. **(I)** Based on sample normality, a two-tailed unpaired t-test was applied for PCS and the Mann-Whitney test for PCG. All values are expressed as mean ±SEM. Legend: PCG, *p*-Cresol glucuronide; PCS, *p*-Cresol sulfate; LC-M, liquid chromatography-mass spectrometry. All values are expressed as mean ±SEM.

**Figure 2.**
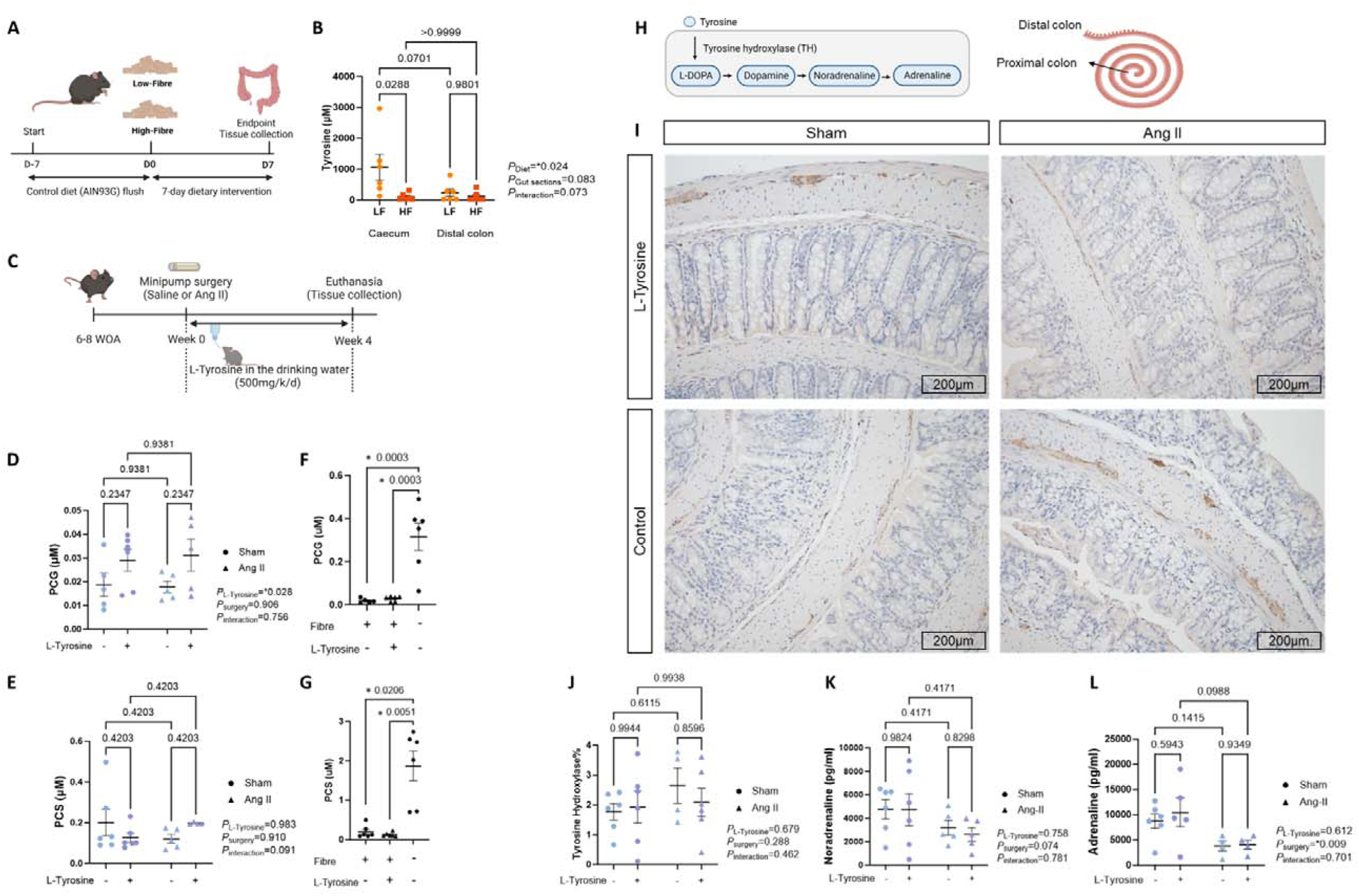
L-Tyrosine supplementation led to an increase in *p*-Cresol-derived metabolites without affecting the host tyrosine biodegradation pathways. **(A)** Experiment design where **(B)** tyrosine level in the caecum and distal colonic contents between mice on a low- or high-fibre diet for a 7-day intervention was measured (2-way ANOVA). **(C)** Study design for L-Tyrosine in hypertension *in vivo*. The relative abundance of **(D)** PCG and **(E)** PCS (μM) in the plasma (Two-way ANOVA adjusted for multiple comparisons; n=5-6/group). The relative abundance of **(F)** PCG and **(G)** PCS (μM) in normotensive mice fed with a standard chow diet with or without L-Tyrosine compared to mice on a low-fibre diet (SF09-028). **(H)** Tyrosine as a substrate for the catecholamine biosynthesis pathway. **(I)** Representative micrographs showing tyrosine hydroxylase (TH) staining (dark brown) in the colon. Scale bar: 200μm. **(J)** The percentage area of the image was positively stained for TH in the colon. The level of catecholamines, **(K)** noradrenaline and **(L)** adrenaline (pg/ml) in the circulation. **(D, E, J-L)** Two-way ANOVA adjusted for multiple comparisons (n=4-6/group). **(F&G)** One-way ANOVA or Kruskal-Wallis test was performed based on data normality (n=6/group). All values are expressed as mean ±SEM.

Next, using an untargeted LC-MS-based approach, we investigated the mouse plasma metabolome in response to different levels of fibre intake and hypertension. The unsupervised hierarchical clustering showed a clear distinction between groups according to diet and surgery (Fig.1B). Thirty-three metabolites were significantly different between low- and high-fibre diets in sham mice (Fig.1C and Supplementary Table 3), and three metabolites in Ang II-treated mice (Supplementary Table 3) after being adjusted for multiple comparisons. Remarkably, PCG was the most significant and higher in abundance in both low-fibre fed sham (*q*=0.006, fold change=10) and Ang II-treated mice (*q*<0.001, fold change=88). Another metabolite closely related to PCG, known as *p*-Cresol sulfate (PCS), was also more abundant in mice on a low-fibre diet (*q*=0.013, fold change=40.9) in the sham animals and similarly was approximately 10-fold greater in the Ang II animals. However, this failed to reach statistical significance once accounting for multiple comparisons. We confirmed the identity of the metabolites identified as PCG and PCS by comparison against authentic standards (Supplementary Fig.3). Both *p*-Cresol and its derived metabolites are uremic toxins that have been reported to be correlated with later CKD stages in humans.^22–24^ Thus, it has been proposed that an accumulation of uremic toxins impairs normal kidney function. However, we did not observe impaired renal function with low-fibre treatment or the dose of Ang II used here.^25^ This suggests that higher levels of PCG may accumulate before renal damage due to low-fibre intake.

Furthermore, we identified that the phenylalanine, tyrosine and tryptophan biosynthesis pathway was the most upregulated metabolic pathway in mice on a low-fibre diet compared to those on a high-fibre diet in both sham and Ang II-treated mice (*q*<0.001, impact factor=1.0, Fig.1D, Supplementary Fig.4A-D and Table 4). Remarkably, the same metabolic pathway was upregulated in Ang II-induced hypertensive versus sham mice (*q*<0.001, impact factor=1.0, Fig.1E and Supplementary Table 5), irrespective of diets (Supplementary Fig.4E-H). Although these pathways are upregulated in the host, this results from an increased bacterial aromatic amino acid catabolism, thereby increasing *p*-Cresol-associated metabolites, potentially associated with low-fibre intake and hypertension.

*p*-Cresol is a metabolite produced and released by anaerobic intestinal bacteria through tyrosine fermentation, and it is the precursor for PCG and PCS.^26,27^ We hypothesised that the increase in *p*-Cresol-associated metabolites in the host was due to fibre deficiency; thus, we investigated the association among fibre intake level, the gut microbiome, and these *p*-Cresol-derived metabolites using Microbiome Multivariable Association with Linear Models (MaAsLin2).^28^ Among the 32 bacteria with statistically significant associations (Supplementary Table 6), nine bacteria were negatively associated with fibre intake level and positively associated with PCG and PCS, showing these microbes were more abundant in a low-fibre-fed microbiome, which in turn were correlated with a higher level of circulating *p*-Cresol derived metabolites (Fig.1F). Bacteria that were correlated with both PCG and PCS showed the same direction, including *Oscillibacter* and *Bilophila* genera. Moreover, we investigated whether there were any changes in *p*-Cresol-related microbial metabolism. In the gut microbiome of a low-fibre diet, we detected a few relevant enzymes that were significantly different (Fig.1G and Supplementary Table 7). For instance, EC2.6.1.57 identified from the *Escherichia* genus, an enzyme converting L-Tyrosine to 4-hydroxyphenylpyruvate and its downstream metabolism leads to *p*-Cresol synthesis, was more prevalent in low-fibre-fed mice. EC4.1.99.2, an enzyme gene involved in L-Tyrosine to phenol metabolism, was also reduced mainly in *Clostridiales bacterium 1_7_47FAA*. Additionally, knowing that *p*-Cresol-associated metabolites were more abundant in mice on a low-fibre diet, these results suggested that most of L-Tyrosine in the colon deviated to the *p*-Cresol generation pathway under a low-fibre diet.

To demonstrate that plasma *p-*Cresol-derived metabolites are not merely associated with the gut microbiota, we used antibiotics to investigate mice with regular versus depleted gut microbiota maintained on a low fibre diet for a week (Fig.1H). The relative abundance of PCG in mice treated with antibiotics was below the detectable range, and PCS was significantly lower compared to mice with normal microbiota (Fig.1I-J). This finding confirms that the production of both *p*-Cresol-associated metabolites under a low-fibre diet depends on the gut microbiota.

### L-Tyrosine supplementation increases p-Cresol-derived metabolites in vivo

Next, we sought to investigate whether fibre intake impacts the level of tyrosine along the large intestine. Importantly, both low- and high-fibre diets had matched levels of tyrosine and other nutrients. However, mice on a low-fibre diet for 7 days (Fig.2A) had increased intestinal tyrosine levels (*P_Diet_*=0.024), particularly in the caecum (*p*=0.029), with higher levels in the caecum than the distal colon (*p*=0.07, Fig.2B). Most amino acid absorption by the host occurs in the small intestine.^29^ Thus, this suggests that the bacteria utilised most of the tyrosine throughout the colon when fibre intake is low. On the other hand, there was no difference in the tyrosine level between caecum and distal colonic contents in mice on a high-fibre diet, suggesting that tyrosine is preferentially utilised in a low-fibre environment.

Therefore, we aimed to investigate whether increased L-Tyrosine intake would lead to an elevated production of *p*-Cresol, subsequently increasing its associated metabolites PCG and PCS (Fig.2C). We measured the circulating levels of PCG and PCS using a targeted metabolomic approach. We found a significant, but modest, increase in PCG in L-Tyrosine-treated animals (*p*=0.028, Fig.2D), independent of the hypertensive status (*P_surgery_*=0.906). The circulating levels of PCS did not change with the L-Tyrosine supplementation among the groups (Fig.2E). Furthermore, we compared the levels of PCS and PCG in normotensive sham mice with or without L-Tyrosine supplementation to mice with a low-fibre intake (from Fig.1H). Mice on a low-fibre diet showed a significant increase in both PCS and PCG levels in the circulation compared to mice fed with a standard chow diet (a max of 4% of crude fibre), supplemented with or without L-Tyrosine (Fig.2F-G). These results strongly suggested that a low-fibre diet, likely due to changes to the gut microbiome, had a much greater impact on circulating PCS and PCG levels than L-Tyrosine supplementation alone.

Tyrosine metabolism is not limited to the gut microbiota. Thus, we investigated whether the host may have directly absorbed the oral L-Tyrosine ingested. L-Tyrosine is a limiting substrate for producing neurotransmitters (Fig.2H), critical players in the sympathetic nervous system in hypertension.^30^ Therefore, we quantified tyrosine hydroxylase (TH) levels, the rate-limiting enzyme that converts L-Tyrosine into L-DOPA, in the colon using immunohistochemistry (Fig.2I). However, there was no difference among the four groups (Fig.2J). We also quantified plasma catecholamine levels to examine whether L-Tyrosine was utilised as a substrate for L-DOPA, promoting the subsequent catecholamine synthesis in the host. Nevertheless, most of the plasma DOPA was below the detectable range (Supplementary Fig.5), reflecting that the circulating level of DOPA was not significantly higher in the L-Tyrosine-treated groups. Ang II-induced hypertensive mice had lower circulating noradrenaline (*P*_surgery_=0.074, Fig.2K) and adrenaline (*P*_surgery_=0.009, Fig.2L) than sham control groups. However, these were independent of L-Tyrosine supplementation. These findings support that most of the L-Tyrosine was likely utilised by the gut microbiota rather than being systemically absorbed and driven to the neurotransmitter biosynthesis pathway to activate the sympathetic nervous system in the host.

### Dietary fibre intake impacts PCG production in humans, which causes an increase in blood pressure

Next, we examined whether findings regarding PCG levels and associated changes to the phenylalanine, tyrosine and tryptophan biosynthesis pathway could be validated in human hypertension. We first used the VicGut cohort (n=68), an untreated Australian cohort with gold-standard (24-hour) BP measurement, where diet, the gut microbiome based on shotgun metagenomic sequencing, and PCG levels were available. Fifty-five per cent of the participants were female, and the cohort mean age was 59.6±7.3 years (Supplementary Table 8). Notably, 61.8% of participants had an inadequate fibre intake, whose PCG level in the circulation was significantly higher than those with an adequate fibre intake (*p*=0.033, Fig.3A), with PCS being borderline significant (*p*=0.058, Fig.3B).

**Figure 3.**
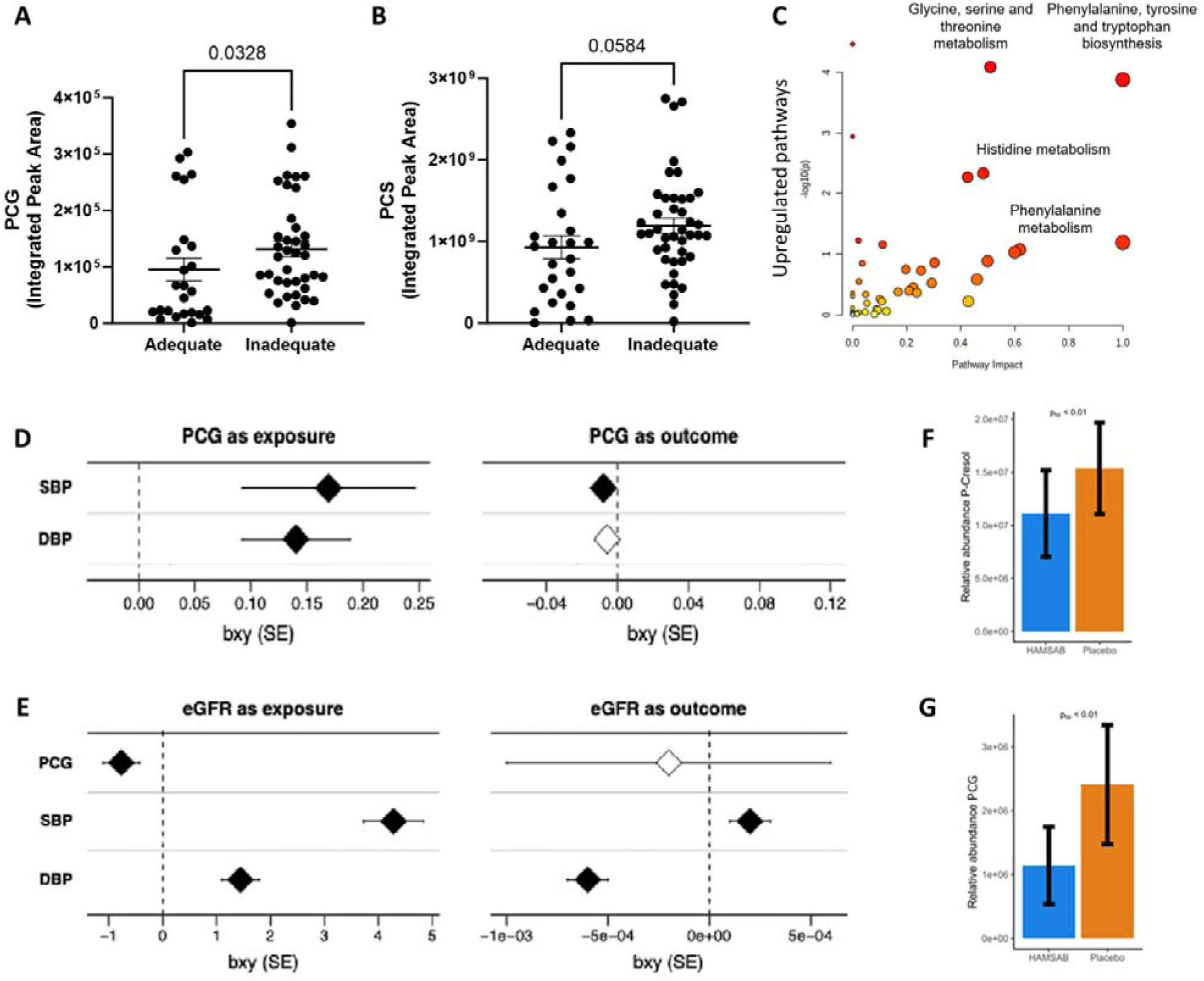
*p*-Cresol glucuronide, derived from an insufficient dietary fibre intake, was causally associated with human hypertension. Human plasma samples were extracted and profiled for their metabolome using LC-MS (VicGut Study). An intake of 25g or greater of dietary fibre per day was considered adequate fibre intake (n=24; inadequate fibre intake n=43). The relative abundance of **(A)** *p*-Cresol glucuronide (PCG) and **(B)** *p*-Cresol sulfate (PCS) in the host metabolome according to fibre intake levels. **(C)** Upregulated pathways in hypertensive versus normotensive subjects (HTN=22; NT=46). Bidirectional Mendelian randomisation **(D)** between PCG and blood pressure traits, and **(E)** between eGFR and PCG or blood pressure metrics. Three weeks of prebiotic HAMSAB intervention were compared to placebo in humans (n=20, crossover design). A moderated t-test (LIMMA) adapted for crossover trial data was performed to assess the effect of HAMSAB on **(F)** *p-*Cresol and **(G)** PCG levels in the circulation. **Legends:** LC-MS, liquid chromatography-mass spectrometry; HTN, hypertension; NT, normotension; PCG: *p-*cresol glucuronide; SBP: systolic blood pressure; DBP: diastolic blood pressure; eGFR: estimated glomerular filtration rate. **(C)** Analyses were performed via MetaboAnalyst using one-way ANOVA, FDR<0.05 and |log2FC|≥1. The pathway enrichment analysis was based on the KEGG Pathway. **(D-E)** The forest plot shows Mendelian randomisation effect results as beta (b_xy_) and standard error (SE) bars. Statistical *p*-value significance is indicated as significant (<0.05) for black diamonds and non-significant (>0.05) for white diamonds.

Association analyses between the human gut microbiome, dietary fibre intake, and the levels of PCG and PCS in the circulation were performed using MaAsLin2 adjusted for age, sex, and BMI. Four bacteria were positively associated with fibre intake, and three bacteria were positively associated with PCG (FDR q<0.05; Supplementary Table 9), with *Ruthenibacterium lactatiformans*, a member of the Oscillospiraceae family, being the most significantly associated with PCG (FDR q=0.001), while none were associated with PCS in the human microbiome. Consistent with our mouse findings, metabolomic pathway analyses revealed that the phenylalanine, tyrosine and tryptophan biosynthesis pathway (*q*=0.003, impact factor=1.0) was the most upregulated in hypertensive participants compared to normotensive participants (Fig.3C, Supplementary Table 10), reflecting microbial activity.

We further validated the relationship among dietary fibre, PCG and hypertension in the TwinsUK cohort, a UK-based cohort study with 1,536 participants, where metabolomic profiling, office BP measurements,^31^ and self-reported use of antihypertensive medication were available. Ninety-two per cent of the participants were female, with a mean age of 55±11.82 years and an average fibre intake of 19.05g/day (Supplementary Table 11). Plasma PCG was associated with dietary fibre intake (*p*=0.011) even after being adjusted for hypertension, age, sex and BMI (Supplementary Table 12). Inversely, PCG was also associated with hypertension (*p*=0.002); however, the significance was lost after adjusting for fibre intake, age, sex, and BMI (Supplementary Table 13), showing that fibre intake is a key driver of PCG levels.

We applied bidirectional Mendelian randomisation (MR) using multiple summary-level genome-wide association study (GWAS) datasets to investigate potential causal relationships between plasma PCG levels and BP.^32^ MR is a well-established approach for inferring causality between modifiable exposures and clinical outcomes.^33^ In forward MR analyses, with PCG as the exposure, genetically predicted PCG levels were significantly and positively associated with higher systolic (b_xy_ [SE]= 0.169 [0.077], *p*=0.029) and diastolic BP (b_xy_ [SE]= 0.140 [0.048], *p*=0.004), based on 23 genetic instruments (Fig.3D and Supplementary Table 14). No evidence of direct causal relationships between PCG and major adverse cardiovascular events was observed (Supplementary Table 14). We performed reverse MR analyses to assess directionality using cardiovascular traits as exposures and PCG as the outcome. Reverse MR detected a very small but significant inverse association of systolic BP with PCG, indicating that higher systolic BP may lead to lower PCG levels (b_xy_ [SE]= −0.008 [0.003], *p*=0.003) (Fig.3D and Supplementary Table 14), suggesting a potential feedback mechanism or confounding biological pathway. This inverse association may reflect pleiotropic effects—such as altered renal or hepatic clearance—rather than a direct influence of BP on PCG production. For instance, elevated BP can initially induce glomerular hyperfiltration, followed by progressive renal impairment.^4^ Consistent with this, we observed that a higher estimated glomerular filtration rate (eGFR) was causally associated with lower PCG levels (b_xy_ [SE]= −0.768 [0.330], *p*=0.020) (Fig.3E and Supplementary Table 14), in line with PCG’s role as a uremic solute cleared by the kidneys. Moreover, bidirectional MR revealed causal links between eGFR and both systolic and diastolic BP (Fig. 3E; Supplementary Table 14), reinforcing the complex interplay between renal function and BP regulation, likely mediated by electrolyte and solute handling. To test whether kidney function confounded the PCG–BP association, we conducted sensitivity analyses by conditioning PCG GWAS summary statistics on eGFR-associated genetic effects (Supplementary Table 15). These revealed that adjusting for eGFR genetic effects led to attenuation and loss of significance of the causal PCG association previously detected for systolic BP (b_xy_ [SE]= 0.140 [0.085], *p*=0.098) and DBP (b_xy_ [SE]= 0.092 [0.052], *p*=0.076) (Fig.6 and Supplementary Table 14). Taken together, these findings suggest that the causal relationship between PCG and BP is at least partially mediated by kidney function, highlighting the importance of renal clearance in modulating circulating microbial metabolites.

Next, by using prebiotic high-amylose maize starch that was chemically acetylated and butyrylated (HAMSAB) to deliver high levels of the SCFAs acetate and butyrate,^34^ we investigated whether a high-fibre diet enriched with SCFAs would rescue the impact of *p*-Cresol and its derived metabolites in a cross-over placebo-controlled double-blind randomised clinical trial. We previously reported that three weeks of HAMSAB intervention led to a 6.1 mmHg reduction in 24-hour systolic BP in patients with untreated essential hypertension relative to the placebo arm.^34^ In a secondary analysis of this trial, we showed that dietary fibre/SCFA intervention significantly decreased the circulating levels of PCG (*q*<0.001, Supplementary Table 16) and *p*-Cresol (*q*=0.023) compared to the placebo arm (Fig.3F-G). However, it did not change the level of PCS (*q*=0.595). Of note, the circulating levels of the amino acid phenylalanine were also lower in the HAMSAB intervention group (*q*<0.024). This amino acid is usually found in protein-containing foods and is generally metabolised into tyrosine in the body.^35^ In contrast, HAMSAB did not change the levels of tyrosine and tryptophan.

### Molecular mechanisms behind p-Cresol-derived metabolites

Finally, we sought to understand how *p*-Cresol-derived metabolites may contribute to BP regulation using a co-expression analysis leveraging whole blood RNA-sequencing and PCG and PCS levels in the TwinsUK cohort (n=933). PCG levels were correlated with 243 genes, and PCS with 231 genes (Fig.4A for negative and Supplementary Fig.7 for positive correlations, all FDR<0.05). The top gene negatively correlated with PCG (and also associated with PCS) was V-set immunoglobulin-domain-containing 4 (*VSIG4).* This gene inhibits the activation of proinflammatory macrophages via the PI3K/Akt–STAT3 pathway.^36^ Ultimately, it reduces inflammatory cytokines associated with hypertension, such as interleukin (IL)-1β and IL-6, via lipopolysaccharide (LPS)-induced macrophage polarisation.^36^ This is relevant as we recently showed that, in the absence of receptors that sense SCFAs, LPS signals via toll-like receptor 4 (TLR4) on macrophages, leading to increased BP.^15^ We then performed pathway analyses and identified that the key pathways negatively associated with PCG levels were related to regulating the inflammatory transcriptional landscape (Fig.4B, Supplementary Table 17). These include genes CXCR2, CD93 and VCP, among others, that play a role in the immune system. Moreover, PCG was associated with the downregulation of endothelial cell development (GO:0001885, q=0.0265, OR=38.95) and endothelial barrier (GO:0061028, adjusted *q*=0.0265, OR=32.71).

**Figure 4.**
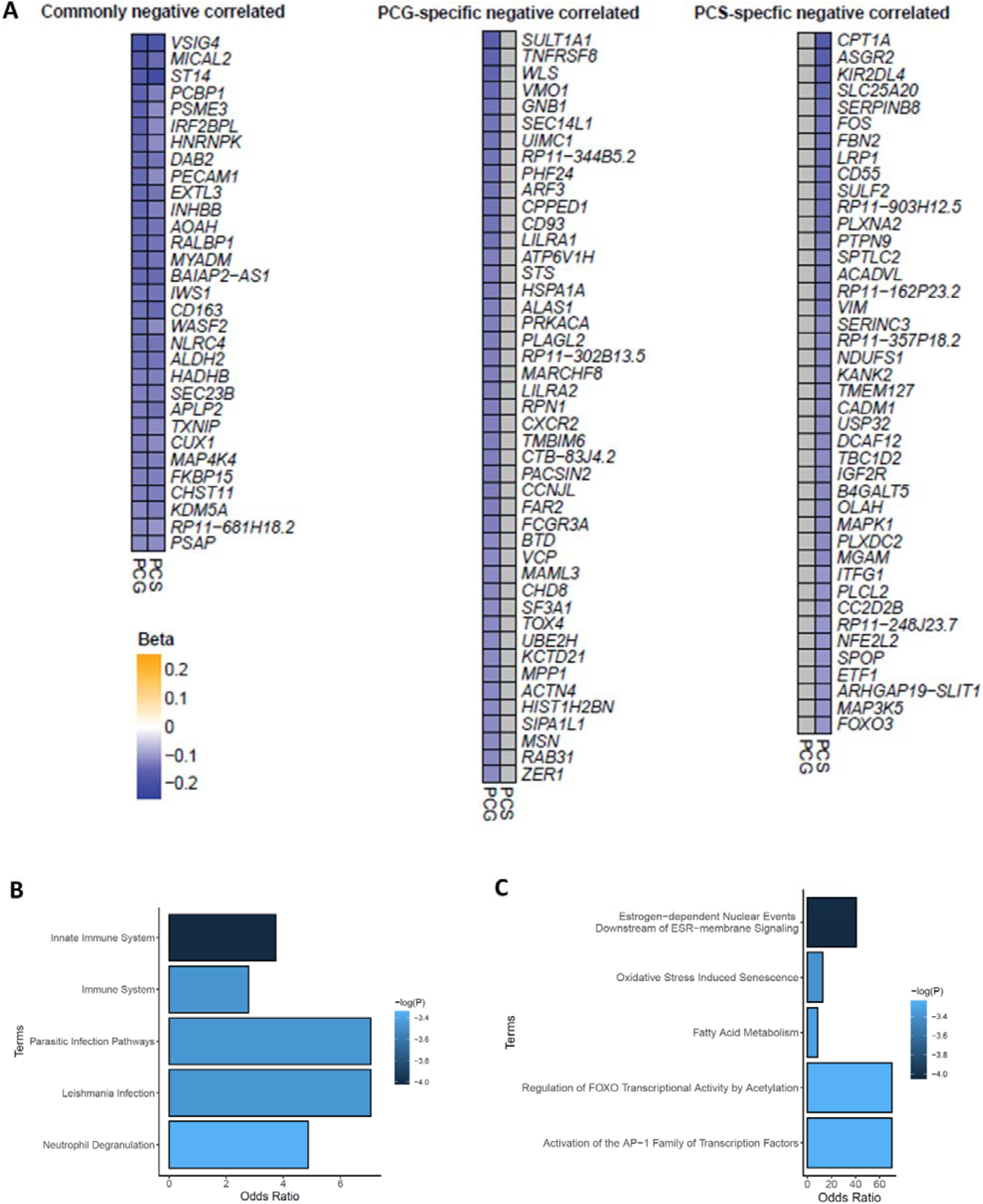
Genes and pathways negatively correlated with *p*-Cresol-derived metabolites. Co-expression analyses were performed between blood RNA-sequencing and *p*-Cresol-derived metabolites in the TwinsUK cohort (n=933). **(A)** Genes negatively correlated to both *p*-Cresol glucuronide (PCG) and *p*-Cresol sulfate (PCS), PCG-specific, and PCS-specific. Pathways negatively correlated with **(B)** PCG and **(C)** PCS, showing the odds ratio of the top 5 differentially regulated pathways. Please see Supplementary Tables 17-18 and Figure 7 for associated data.

In contrast, PCS was negatively associated with pathways related to oxidative stress-induced senescence, fatty acid metabolism, and regulation of FOXO transcriptional activity by acetylation (Fig.4C, Supplementary Table 18) – this may be related to the lower levels of SCFAs, which can be converted to acetyl-CoA, providing the acetyl group for acetylation.^37^ No pathways were significantly upregulated in association with PCG or PCS.

## Discussion

Most countries are not reaching the minimum recommended fibre intake.^19,20^ This study identified a novel host–microbiota–metabolite axis explaining how insufficient dietary fibre intake elevates BP. Under fibre-deficient conditions, microbial communities reprogrammed their metabolic preferences toward tyrosine fermentation, substantially increasing circulating *p*-Cresol-associated metabolites. Among these, only PCG was enriched in experimental hypertension, implicating it as a potential mechanistic mediator. The coordinated upregulation of phenylalanine, tyrosine, and tryptophan biosynthesis in both human and murine hypertension supports the notion of heightened amino acid demand by the microbiota, reflected in the host plasma, in response to insufficient fibre intake. Genetic approaches demonstrated that elevated PCG contributes causally to BP elevation in a kidney function– dependent manner. Notably, dietary fibre enriched with SCFAs supplementation in hypertensive individuals reduced PCG levels, highlighting its therapeutic potential to modulate and reduce uremic toxin load and cardiovascular risk. Finally, our findings reveal a transcriptional signature of PCG-mediated immune and endothelial modulation, providing mechanistic insight into its links with hypertension.

Herein, we confirmed fibre intake was a potent modulator of the gut microbiome, consistent with previous literature from our team^12,34,38,39^ and others in mice^40–42^ and humans.^43–46^ The effect of dietary fibre on the gut microbiome could impact microbial metabolism, which, in turn, alters the metabolome in the host. We found that Oscillospiraceae family was more enriched in low-fibre-fed mice, and surprisingly, was also positively associated with human PCG levels. The abundance of *Oscillibacter* was previously seen in mice fed a high saturated fat diet, where it was inversely associated with the mRNA expression of zonula occludens-1 (*Zo-1*), an essential gut integrity protein.^47^ Surprisingly, this bacterium was positively associated with four uremic metabolites, including PCS, PCG, trimethylamine N-oxide (TMAO) and indoxyl sulfate (IS) in the serum, and correlated with increased kidney function deterioration in patients with moderate to severe CKD.^48^ In the VicGut cohort, *Ruthenibacterium lactatiformans*, a member of the Oscillospiraceae family previously reported to be positively associated with an increased cardiovascular risk,^49^ was positively associated with PCG. Notably, under conditions of insufficient fibre intake, the levels of PCG and PCS were higher in the circulation in both mice and humans in the VicGut study; this was further validated in the TwinsUK cohort. The precursor of these two metabolites is *p*-Cresol, mainly derived from the biotransformation of undigested tyrosine-containing food in the large intestine.^35^ Some anaerobic intestinal bacteria metabolise tyrosine into *p*-Cresol and release it as a by-product.^26,27^ As a result, *p*-Cresol crosses through the colonic mucosa and enters the circulation, gaining its entrance to the liver. Antibiotics reduced the levels of PCG and PCS in the circulation to a barely detectable level, demonstrating that the gut microbiota has a crucial role in regulating the levels of *p*-Cresol-associated metabolites in the host.

On a low-fibre diet, mice had higher caecal tyrosine levels than in the distal colon, showing that the bacteria likely fermented tyrosine from the undigested food mass throughout the large intestine. In contrast, the level of tyrosine in the caecum and distal colon remained the same in mice on a high-fibre diet. Mouse metagenome sequencing revealed that the enzyme involved in the metabolism of L-Tyrosine to 4-hydroxyphenylpyruvate (EC2.6.1.57), an upstream metabolism step of the *p*-Cresol synthesis,^50^ was more active in the microbiome of mice fed a low-fibre diet. Our findings suggest that under fibre deprivation, the gut microbiota switched their resource preference, fermented tyrosine and increased its metabolism, eventually leading to higher levels of PCG and PCS in the circulation.

Gut microbiota-derived metabolites PCG and PCS have been previously associated with incident risks of major adverse cardiovascular events and all-cause mortality;^51^ however, a key question remained whether they contribute towards cardiovascular risk or are merely associated with it. Our MR results indicated that PCG levels were causally associated with higher BP. This was, however, mediated by renal function, likely reflecting renal impairment in chronic hypertension. Uremic toxins are found to induce endothelial dysfunction in CKD patients through different mechanisms. For example, some uremic toxins were reported to increase oxidative stress and pro-inflammatory and pro-thrombotic proteins.^52^ A recent study reported that the uremic toxins PCS and IS enhanced the development of thrombosis.^53^ The concept was tested using germ-free mice colonised with bacteria engineered to produce indole (a precursor for IS) and *p*-Cresol, which resulted in a pro-thrombotic phenotype.^53^ Remarkably, in our randomised clinical trial, the three weeks of prebiotic intervention reduced PCG and *p*-Cresol levels in the circulation in the HAMSAB arm compared to the placebo arm. Similarly, plasma *p*-Cresol was decreased in CKD patients with dietary fibre intervention (pea hull combined with or without inulin, a type of fermentable fibre).^54^ Additionally, we identified that reduced PCG levels were correlated with the inflammatory transcriptional landscape in the TwinsUK cohort. These findings showed the potential of dietary fibre to reduce uremic toxins and, promisingly, reduce inflammation by shifting the food resources available to the gut microbiota.

We acknowledge that this study has some limitations. All the mice studied were male; however, both sexes were present in the three independent human studies. PCG is a uremic toxin and may induce renal damage. However, the mouse strain used, C57BL/6J, is resistant to kidney disease-associated pathologies,^55,56^ thus, we did not observe renal damage with low-fibre intake. This study sequenced the mouse gut microbiome from caecal samples, where fibre fermentation occurs prominently. However, amino acid fermentation is mainly active in the distal colon.^35^ Therefore, future studies studying low-fibre intake should explore the gut microbiome in the distal colon, where most protein fermentation occurs. Future studies could also investigate the ability of *p*-Cresol inhibitors (such as 4-Hydroxyphenylacetonitrile^57^) to reduce PCG and its associated cardiovascular risk, or block renal clearance. A limitation of this study is that MR relies on the validity of genetic instruments, which may be affected by horizontal pleiotropy or confounding pathways— particularly in complex traits like BP and kidney function—potentially biasing causal estimates. To experimentally validate the MR findings, we explored direct delivery of PCG to mice. However, rapid clearance of PCG in healthy animals, combined with the high infusion volume required for sustained administration via osmotic minipumps, exceeded permissible dosing limits and rendered the experiments unfeasible.

In conclusion, using a combination of experimental and human studies, our findings uncover a previously unrecognised mechanistic link between insufficient dietary fibre intake, microbial metabolism, and the pathogenesis of hypertension. Under fibre-deprived conditions, the gut microbiota shifts resource preference and ferments tyrosine into *p*-Cresol, increasing PCG in the host circulation. This metabolite increases human BP, particularly when renal function is decreased, likely via immune and endothelial mechanisms. Remarkably, dietary fibre intervention in humans reduced PCG and *p*-Cresol, highlighting a tractable strategy for modulating circulating uremic toxins and mitigating cardiovascular risk.

## Methods

### Animal model and dietary intervention

All experimental protocols were approved by the Alfred Medical Research and Education Precinct Animal Experimentation Ethics Committee (E/1626/2016/B). Animals used in this study were described in detail elsewhere.^12^ Briefly, 3-week-old male wild-type (WT) C57BL/6J mice were randomly allocated to a diet lacking resistant starches (referred to as ‘low fibre’, SF09-028, Specialty Feeds) or a diet high in resistant starches (all carbohydrates replaced with resistant starches, referred to as ‘high fibre’, SF11-025, Specialty Feeds), eight animal per group. Both diets contained a similar amount of protein, carbohydrates, lipids, vitamins, sodium, potassium, and sodium-to-potassium ratio. Mice received the diets from three weeks before minipump surgery until the end of the study. Six-week-old mice from each group were further randomised for surgical implantation of a minipump containing either saline (0.9%, called sham) as control or a low dose of Ang II (0.25 mg/kg/d) for four weeks (n=4/group, total 16 mice). Animals were monitored and weighed regularly. At the endpoint, mice were euthanised by CO_2_ asphyxiation, EDTA plasma and caecum content were collected and stored at −80°C. Metabolomic profiling of plasma samples and microbiome sequencing of caecum content samples were performed, both of which are described below.

### Caecal DNA extraction, library preparation and sequencing

Mouse caecal DNA and human faecal DNA were extracted using the DNeasy PowerSoil DNA isolation kit (Qiagen). The V4-V5 region of the bacterial 16S rRNA was amplified by PCR using 20 ng of sample DNA was mixed with Platinum Hot Start PCR master mix (ThermoFisher Scientific), 515F and 926R primers (Bioneer) and amplified in a Veriti Thermal Cycler (ThermoFisher Scientific). The quality and quantity of the PCR product were assessed in a MultiNA MCE-202 Microchip Electrophoresis System (Shimadzu) using the DNA-2500 kit (Shimadzu), and 240 ng of PCR product per sample was pooled and cleaned using the PureLink PCR Purification kit (ThermoFisher Scientific). The product was then sequenced in an Illumina MiSeq sequencer (300 bp paired-end reads).

We also performed shotgun metagenome sequencing (GENEWIZ) to investigate the differences in the gut microbiome between low- and high-fibre intake. One to two biologically independent caecal DNA samples from the same animal cohort were pooled together. DNA was fragmented by acoustic disruption using Covaris S220 and then underwent end repair, dA-tailing, adapter ligation (P7 and P5 primers) and purification. The purified DNA was further selected for the right size before PCR amplification for library construction. The preliminary quantification and dilution of the library is performed using Qubit3.0, and then Agilent 2100 was used to determine the insert size and nucleic acid concentration of the resulting library sample. The effective concentration of each sample library in the library mixture was determined by quantitative PCR (qPCR) before sequencing to ensure the accuracy of the sample concentration and the reliability of the sequencing data. The product was then sequenced on an Illumina NovaSeq instrument with 30 million/reads per sample.

### Bioinformatic analyses of the faecal microbiome

#### a) 16s rRNA microbiome data analyses

Sequence reads from samples were first analysed using the Quantitative Insights Into Microbial Ecology version 2 (QIIME2) framework.^58^ Forward and reverse reads were first truncated at base number 225 for forward reads and base number 160 for reverse reads, then were denoised, merged, and chimera filtered using the DADA2 plugin,^59^ resulting in an operational taxonomic units (OTUs) table (via q2-dada2) aligned with the SILVA database (version 138)^60^ with resolution at the single nucleotide level. A phylogeny tree was then created using fasttree2^61^ from mafft-aligned.^62^ OTUs and was subsequently subsampled without replacement to 28,160 counts per sample, with an average of 28,160 counts per sample (via q2-alignment and q2-phylogeny). β-diversity metrics were generated from the rarefied samples (via q2-diversity), reported here as unweighted and weighted Unifrac metrics along with associated Principal Coordinate Analysis (PCoA) distance tables. The distance metrics from PCoA and β-diversity were evaluated using the non-parametric Permutational Multivariate Analysis of Variance (PERMANOVA) to test for differences in microbial community composition between treatment groups. α-diversity metrics, including Chao1 and Shannon indices, were analysed using MicrobiomeAnalyst (version 2.0).^63,64^ edgeR using P-value adjusted with false discovery rate (FDR, *q*<0.05) was performed to compare the differences between low- and high-fibre on MicrobiomeAnalyst. In addition, the R package Microbiome Multivariable Association with Linear Models (MaAsLin2)^28^ was used to correlate individual bacteria with variables such as fibre intake and specific metabolites.

#### b) Shotgun metagenome sequencing

FASTQ files were first inspected with FastQC (version 0.11.9).^65^ Trimmomatic (version 0.38) was then used with the paired-end flag for Illumina adapter trimming, leading and trailing removal of bases with a quality score below 3, and for cutting each read using a 4-base wide sliding window where the average per-base quality dropped below 15.^66^ Contig and scaffold assembly were conducted using metaSPAdes (version 3.13.1) with 96 GB of RAM and 64 threads.^67^ Separately, FASTQ files were passed through HumanN2, supported by Bowtie2 (version 2.3.5) and the TBB library (version 20180312oss).^68,69^ HumanN2 was run with the protein-database, nucleotide-database, and metaphlan-options flags, pointing to the UniRef, chocoPhlan, and MetaPhlAn databases downloaded with the included humann_databases utility script. The humann_rename_table utility script was used to reattach full gene and pathway names, and the humann_renorm_table script was used to convert reads per kilobase into normalised copies per million counts. The gene count outputs were further condensed by grouping genes by level-4 Enzyme Commission categories using the humann_regroup_table utility script. Output files were then passed to R (version 4.0.3), where gene counts, level-4 Enzyme Commission category-grouped genes, and pathway abundance counts were analysed for differential expression between diets using the limma package (version 3.46.0), supported by the Tidyverse package (version 1.3.1).^70–73^ Differential abundance of taxa was analysed using the ALDEx2 package (version 1.22.0) using the main Aldex function with “zero” as the set of features to retain as the denominator for the subsequent geometric mean calculation.^74^ PCoA plots and volcano plots were then created using ggplot2 (version 3.3.5).^75^ Linear discriminant analysis (LDA) effect size (LEfSe) analyses were performed through the Galaxy platform.^76^

### Plasma extraction and metabolomics profiling

Metabolites were extracted from 25 µl of EDTA plasma by the addition of 200 µl of 1:1 methanol/acetonitrile with a mixture of CHAPS, CAPS, PIPES and TRIS at 1 µM each. Samples were briefly vortexed and then mixing for 20 minutes on a vibrating mixer at 4°C before centrifugation (20,000 ×g, 15 minutes at 4°C). The supernatant was transferred to an sample vial with glass insert for LC-MS analysis.

For LC-MS analysis, samples were analysed by hydrophilic interaction liquid chromatography coupled to high-resolution mass spectrometry (LC-MS).^77^ In brief, we utilized a polymeric ZIC-pHILIC 5 µm, 150 x 4.6 mm column (Merck, Australia) at 25°C with a gradient elution of 20 mM ammonium carbonate (A) and acetonitrile (B) (linear gradient time-%B as follows: 0 min-80%, 15 min-50%, 18 min-5%, 21 min-5%, 24 min-80%, 32 min-80%) on a Dionex RSLC3000 UHPLC (Thermo). The flow rate was maintained at 300 µL min^-1^. Samples were kept at 4°C in the autosampler and 10 µL injected for analysis. Mass spectrometry was performed on a Q-Exactive Plus Orbitrap (Thermo Fisher Scientific, Australia) in MS1 polarity switching mode. The instrument was operated at 35,000 resolution with the following conditions: Electrospray ionization voltage was 4 kV in positive and −3.5 kV in negative mode, capillary temperature = 300 °C; sheath gas = 50; Aux gas = 20; sweep gas = 2; probe temp = 120°C). Acquired LC-MS data was processed in an untargeted fashion using opensource software IDEOM, which initially used msConvert (ProteoWizard)^78^ to convert raw LC-MS files to mzXML format and XCMS to pick peaks to convert to.peakML files.^79^ Mzmatch was subsequently used for sample alignment and filtering.^80^ This workflow results in a dataset in which the majority of the metabolites are only putatively identified and consequently care must be taken in interpreting that data biologically. To aid accurate metabolite identification, a standard library of ∼500 metabolites was analysed before sample testing and accurate retention time for each standard was recorded. This standard library also forms the basis of a retention time prediction model used to provide putative identification of metabolites not contained within the standard library.^81^ Additionally, in the case of PCG and PCS the identity of the metabolites were subsequently verified on the basis of their retention time by comparison of authentic standards against representative samples (Supplementary Fig. 3).

### Metabolomics data analyses

#### a) Statistical analyses

All statistical methods were conducted through MetaboAnalyst platform (version 5.0), which provides tools for metabolomic data analyses.^82^ Missing values were imputed with one-fifth of the lowest detected value.^82^ Although peak intensities were recorded as continuous and numeric values, they were bounded between zero and positive infinity. Thus, to avoid violating assumptions for statistical analyses, such as the Student’s t-test, a logarithm base 10 was used to transform the intensity values. After quality checks, unsupervised learning techniques were applied to examine whether there was a separation between groups based on their metabolomics. A hierarchical clustering (presented here as a heatmap) is an unsupervised method that does not consider data labelling; thus, samples are grouped based on their metabolomic profiles, not on the group. The parameters for building heatmaps included one-way analysis of variance (ANOVA) and post-hoc (Fisher’s least significant difference) analyses (*p*<0.05) to detect the top 50 compounds that vary amongst the samples, and Euclidean distance measure and Ward clustering algorithm. Data were analysed according to fibre intake and hypertensive status. The Student’s t-test was chosen to compare the mean intensity of metabolites between each pair of study groups. All *p* values were adjusted for FDR to control the number of false positives, and *q*<0.05 was considered statistically significant.

#### b) Pathway analyses

To explore the relevant biological pathways involved in BP and fibre intake, all putative metabolites identified by the IDEOM analysis (in their un-transformed values) were considered,^79^ and the fold change for each pair of study groups was calculated. Each group was analysed independently via the MetaboAnalyst platform (version 5.0). Mouse and human data were analysed using the *Mus musculus* and the *Homo sapiens* pathway library, respectively. The hypergeometric test was used as the method for over-representation analysis, which examines whether metabolites involved in a pathway were enriched more than expected by chance within the dataset. Relative betweenness centrality was chosen for topological analysis (visualisation output).

### Antibiotics study

All experimental procedures were approved by the Monash Animal Ethics Committee (AEC approval 27929). Six to eight weeks of age (WOA) male C57BL/6J WT mice received a week of the control diet (AIN93G) to standardise the gut microbiome baseline. Then, they were randomised and placed under either normal drinking water or water treated with an antibiotic cocktail of enrofloxacin (10mg/kg body weight/day) and amoxicillin with clavulanic acid (50mg/kg body weight/day) to reduce the gut bacteria load until the end of the experiment (n=6 per group); bacteria load results reported in.^83^ From the seventh day of antibiotic treatment, mice were fed a low-fibre diet (SF09-028) until the experiment endpoint. Whole blood was collected via cardiac puncture into EDTA tubes, and plasma was isolated. A targeted LC-MS reverse phase method was used to detect the relative abundance level of *p*-Cresol glucuronide (PCG) and PCS in the plasma samples (details below).

### Metabolomic profiling

Plasma samples were allowed to thaw at 4°C for 1 hour. 75μL of ice-cold acetonitrile was added to 25μL of plasma and the mixture was vortexed briefly before being mixed on a PVC 3000 vortex mixer (1500 rpm, 1 centrifugation sec, hard, vortex 20 sec, 60 cycles) at 4°C (cold room). Samples were then subjected to centrifugation at 22,000 rcf for 10 minutes at 4°C. Then, 20μL of each supernatant was transferred to a sample insert before adding an additional 55μL of H_2_O (bringing the solvent composition to approximately 20% acetonitrile) for LC-MS analysis. A Dionex RSLC3000 UHPLC coupled to a Q-Exactive Plus Orbitrap MS (Thermo) was used for analysis. Samples were analysed by reverse phase chromatography, specifically a Zorbax Eclipse Plus C18 Rapid Resolution HD 2.1 x 100mm 1.8 micron column (Agilent, Australia). A gradient elution of 0.1% formic acid (A) and 0.1% formic acid in acetonitrile (B) (linear gradient time-%B: 0 min-2%, 15 min-100%, 19 min-100%, 21 min-2%, 26 min-2% was utilised at 40 °C. The flow rate was maintained at 300μL/min. Samples were kept in the autosampler (6°C), and 10μL was injected for analysis. MS was performed in negative mode, alternating between full scan MS at 70,000 resolution and MS/MS for the compounds of interest (PRM mode) at a resolution of 17,500. ESI conditions were as follows: 3.5kV spray voltage; 300°C capillary temperature; 50 sheath gas flow rate; 20 auxiliary gas flow rate; 2 sweep gas; 120°C probe temperature. Extracted ion peak integration was performed on the product ion (PCG m/z = 107.05024 Th, 5.08 min; PCS m/z = 107.05024 Th, 5.86 min; PCS-d4 m/z = 111.07500, 5.83 min) using TraceFinder 4.1 (Thermo Scientific, Australia). Calibration curves were constructed using plasma from a sham control mouse known from previous experiments to have a low endogenous concentration of PCG and PCS. The PCS curve was constructed using a deuterium labelled version of PCS, (PCS-d4).

### Tyrosine detection by colorimetric kit

Tyrosine levels were measured in the caecum and distal colon samples using the colorimetric kit (Sigma-Aldrich Tyrosine Assay Kit, mak219) on WT animals fed on low-(SF09-028) or high-(SF11-025) fibre diet for 7 days (AEC approval 27929). Caecum content (25mg) was weighed and homogenised in 500μl of distilled water (1:20 w:v ratio), centrifuged at 8,000*g* for 5 min at 4°C. The supernatant was filtered through a 40-μm cell strainer into 50ml Falcon tubes, and the filtrate was transferred into a 10-kDa spin column (Abcam) and proceeded the following steps according to the manufacturer’s instructions.

### L-Tyrosine in vivo

#### a) Animal model and intervention

Male C57BL/6J WT mice at 6–8 weeks old were randomised and underwent minipump surgery to receive either saline (0.9% sodium chloride, called sham) as control or Ang II (0.5mg/kg/day) to elicit a hypertensive response (n=8/group). Following the surgery, mice received either regular water or L-Tyrosine (500mg/kg/day) in the drinking water (n=4/group); the latter was freshly prepared and changed at least twice per week, as recently published.^84^ The mice had standard housing and husbandry throughout the experiment in a specific pathogen-free facility. We had two independent animal cohorts applying the same animal model and interventions. Each cohort consisted of 16 mice in total, with four individuals per treatment group. All experimental procedures were approved by the Monash Animal Ethics Committee (AEC approval 28316). Plasma was collected at the animal endpoint and run for targeted metabolomics as mentioned above.

#### b) Histological analyses

Paraffin-embedded Swiss-rolled colon tissue samples were cut into 4μm-thick sections and used for immunohistochemistry to analyse tyrosine hydroxylase (TH) in the gut. All staining methods were performed according to the manufacturer’s protocol. Briefly, TH primary antibody (unconjugated, Merck, AB152) at 1:500 dilution and Rabbit-specific HRP/DAB Detection Kit (Abcam 64261) were used. Images were acquired using a Nikon A1R confocal microscope at 400× magnification. Analysis was performed blinded using ImageJ software with an automated macro (developed by Chad Johnson).

#### c) Catecholamine detection by high-performance LC analysis

Plasma catecholamines were extracted and quantified as previously described.^85,86^ Briefly, plasma (50-200μl) was placed in 0.5ml of 0.4M perchloric acid containing 0.01% EDTA with 3,4-dihydroxybenzylamine. Samples were centrifuged, and the supernatant was collected. Catecholamines were extracted from the supernatant using alumina absorption, separated by high-performance liquid chromatography, and concentration was quantified by colourimetric detection as described elsewhere.^85,87^

#### d) Statistical analyses

Statistical analysis and graphing were performed using GraphPad Prism (versions 9 to 10.0.2). The ROUT method (Q=1%) was performed to identify outliers. The normality of data was determined using Shapiro–Wilk’s normality test. For normally distributed data, a two-tail unpaired t-test was used to compare between two groups. For non-parametric data, the Mann-Whitney test was performed. For a 2×2 data, two-way ANOVA (adjusted with Tukey’s false discovery rate for multiple comparisons) was used to compare the data between groups. Data are presented as mean ± standard error of the mean (SEM), and those with a *P* (adjusted for multiple comparisons when there were more than two groups) < 0.05 were considered significant.

### Victoria Gut Cohort

Seventy volunteers were recruited for the Victoria Gut (VicGut) cohort; the main characteristics of this cohort were previously described in detail elsewhere.^18^ This cross-sectional study included participants from two sites: 40 were recruited from a metropolitan clinic (Alfred Hospital, Melbourne, Victoria, Australia), and 30 were from a regional clinic (Shepparton, Victoria, Australia). The study complied with the Declaration of Helsinki and was approved by the human research ethics committee of the Alfred Hospital (approval 415/16). All subjects gave informed consent and the study was registered under ACTRN12620000958987.

#### a) Selection criteria

Inclusion criteria included being 40-70 years of age, either sex, having a body mass index (BMI) of 18.5 to 30, and not receiving BP-lowering medication. Exclusion criteria included having any gastrointestinal disease (including a history of intestinal surgery, inflammatory bowel disease, celiac disease, lactose intolerance, chronic pancreatitis, or other malabsorption disorder), having type 1 or 2 diabetes, chronic kidney disease, or having used probiotics or antibiotics in the past three months.

#### b) Blood pressure measurement

Office BP was measured 2-3 times using an automatic BP monitor (OMRON) by trained staff. Participants were then fitted with an ambulatory (24-hour) BP monitor device. Hypertension was diagnosed when 24-hour systolic BP and/or diastolic BP was greater than or equal to 130 and/or 80 mmHg, respectively, according to the European and Australian guidelines.

#### c) Food frequency questionnaires

Dietary intake over a 12-month period was assessed using the Dietary Questionnaire for Epidemiological Studies (DQES) version 3.2, a self-administered and validated food frequency questionnaire (FFQ) developed by the Cancer Council Victoria, which reflects the dietary intake of the Australian population. A total of 67 participants completed the FFQ. Participants were classified as having inadequate fibre intake (referred to as “low-fibre intake”) when their total dietary fibre was less than 25g per day, or adequate fibre intake (referred to as “high-fibre intake”) when the total dietary fibre was greater than or equal to 25g per day. This threshold was devised based on the findings from a meta-analysis showing that a minimum of 25g/day decreases the incidence of cardiovascular diseases.^7^

#### d) Faecal DNA extraction, library preparation and sequencing (as above)

#### e) Microbiome metagenome bioinformatic analyses (as above)

#### f) Plasma extraction, metabolomic profiling and analyses (as above)

Methods as described above (total n=68). Blood samples were collected from the participants for metabolomic profiling; however, participant #93 was not included in the blood collection. Sample #94 was excluded on the basis of a low median peak intensity and low internal standard signal indicating an LC-MS fault (most likely a skipped injection); thus, a total of 68 metabolomic samples were available in this cohort.

#### g) Statistical analyses

Data were first screened for outlier identification using the ROUT (Q=1%) and assessed for their normality distribution using the Shapiro-Wilk test. The Mann-Whitney test was performed on non-parametric data to assess the levels of fibre intake and the *p*-Cresol-derived metabolites (PCG and PCS) in the circulation.

### TwinsUK Cohort

Replication was conducted in individuals from the TwinsUK cohort, a deeply phenotyped adult twin registry comprising over 16,000 participants with extensive phenotypic and biological data.^88^ TwinsUK participants are predominantly middle-aged females with equal proportions of monozygotic and dizygotic pairs. Ethical approval for the TwinsUK study was granted by the St Thomas’ Hospital Research Ethics Committee, and all participants provided written informed consent (REC ref EC04/015).

#### a) Dietary data

Dietary data in TwinsUK were collected using a validated 131-item semi-quantitative food frequency questionnaire (FFQ) originally developed for the EPIC-Norfolk study.^89^ Fibre intake was estimated from the FFQ using the FETA software.^89,90^ and adjusted for energy intake using the residual method.^91^

#### b) Metabolomics

*p*-cresol metabolites, *p*-cresol sulfate, and *p*-cresol glucuronide were measured by Metabolon inc.(Durham, USA) using an untargeted LC/MS and GC/MS platform from fasted serum samples as previously described.^31,92,93^ Samples were stored at −80°C until processing. After methanol precipitation and centrifugation (Glen Mills GenoGrinder 2000), the extract was split into five fractions: aliquots (1) and (2) were analysed under acidic positive ion conditions optimised for hydrophilic and hydrophobic compounds, respectively; aliquot (3) under basic negative ion conditions using a dedicated C18 column; aliquot (4) using negative ionisation after HILIC separation; and aliquot (5) was reserved as backup. Controls included extracted water (process blanks) and a standard cocktail spiked into each sample to monitor instrument performance and aid chromatographic alignment. Instrument variability was assessed via median relative standard deviation (RSD) of internal standards, while process variability was determined from the median RSD of metabolites found in all pooled technical replicates. Samples and controls were randomised across the run.

*Compound identification* Metabolites were identified by comparison of the ion features in the experimental samples to a reference library of chemical standard entries that included retention time/index, molecular weight (m/z), and MS spectra. Identification of known chemical entities is based on comparison across all three features to metabolomic library entries of purified standards.

*Metabolite quantification and normalisation* Peaks were quantified using area-under-the-curve. Raw area counts for each metabolite in each sample were normalised to correct for variation resulting from instrument inter-day tuning differences by the median value for each run-day, therefore, setting the medians to 1.0 for each run. This preserved variation between samples but allowed metabolites of widely different raw peak areas to be compared on a similar graphical scale.

*QC* To remove batch variability, for each metabolite, the values in the experimental samples were divided by the median of those samples in each instrument batch, giving each batch and thus the metabolite a median of one. Missing values were imputed, using the minimum value across all batches in the median scaled data.

#### c) p-cresol metabolites, fibre, and hypertension analysis

This analysis included 1,536 participants with metabolomic profiling, office BP measurements,^31^ and self-reported use of antihypertensive medication. Linear mixed-effects models (‘lme4’ R package) were employed, adjusted for age, sex, BMI, and the presence or absence of hypertension as fixed effects while accounting for family-relatedness as a random effect.

#### d) Blood sample extraction and RNA-sequencing

MultiMuTHER, a longitudinal sub-study of TwinsUK, enrolled 335 female participants (148 MZ twin pairs, 166 DZ twins, 21 singletons). Whole-blood RNA and serum samples were collected at three time points over a median 6-year interval. Extracted RNA was prepared with the Illumina TruSeq kit and sequenced on the Illumina HiSeq 2000. 49 bp paired-end reads were aligned to GRCh38 using STAR v2.6.1a^94^ (MAPQ ≥ 255). Gene-level counts per million (CPM) were generated with the quan module of QTLtools,^95^ retaining genes expressed at ≥5 counts in ≥25 % of samples. Between-sample normalisation used trimmed-mean-of-M-values (TMM),^96^ after which each gene was rank-based inverse-normal transformed (rntransform function from the GenABEL package).^97^

#### e) RNA-sequencing and pathway co-expression analyses

UHPLC-MS/MS profiling (Metabolon Inc.) was conducted on 993 serum samples across the 335 study participants. For each run-day, metabolite intensities were median-scaled to 1, and missing values were imputed using random-forest non-parametric imputation (R package missForest.^98^

Inverse-normalised metabolite concentrations were regressed on residualised gene-expression values using linear mixed-effects models (lme4 package in R 3.6).^99^ Expression residuals were first adjusted for RNA sequencing technical covariates transcript integrity number (TIN), insert size, GC content, RNA integrity number (RIN), library preparation date, sequencing date and primer index. Metabolite-gene expression GEAS mixed effects model fixed effect covariates were age at visit, BMI at visit, time-of-day, seasonality, and metabolite-specific year of visit effects. Random effects included were Metabolon sample processing plate, family ID, zygosity and participant ID. One degree of freedom likelihood-ratio tests were used to compare full models to null models omitting the tested gene. To adjust for multiple testing, we applied a study-wide Benjamini–Hochberg FDR, and retained gene– metabolite associated pairs with FDR < 0.05.

### Mendelian Randomisation Analysis

To test for causal effects between PCG and cardiovascular outcomes, we performed a generalised summary data-based Mendelian Randomisation (MR) analysis using GSMR2 (implemented in GCTA v1.94.1, https://yanglab.westlake.edu.cn/software/gsmr/).^32,100^ Based on a two-sample, multi-SNP model, this method uses genetic instruments as proxies for an environmental modifiable exposure (in this case, PCG levels) to test for a putative causative association with an outcome, employing genome-wide association study (GWAS) summary-level data from two independent studies. MR is based on the principle that genetic instruments are unlikely to be confounded in the way that direct measures of the exposure might be in observational studies.^32^ In addition, we also applied the HEIDI (heterogeneity in the dependent instrument) outlier filtering method to detect and remove SNPs that have potential horizontal pleiotropic effects on both the exposure and outcome (HEIDI-outlier p-value < 0.01). Notably, pleiotropy is a key factor that can bias estimates and inflate test statistics in MR analysis.^32^

The cardiovascular outcomes of interest included systolic BP (SBP), diastolic BP (DBP), and major adverse cardiovascular events, particularly acute coronary syndrome (ACS), heart failure and ischaemic stroke. Data on the effects of genetic variants on exposure and cardiovascular outcomes were retrieved from large studies available at the GWAS Catalog (https://www.ebi.ac.uk/gwas/). Full GWAS summary statistics on individuals of European genetic ancestry were publicly available for all cardiovascular outcomes, with sample sizes ranging from 446,696 to 1,028,900 (Supplementary Table 19). For PCG, GWAS results on 8,066 European individuals from the NIHR UK Bioresource cohort, adjusted for age, sex, and BMI, were accessed upon request (through https://hidrive.ionos.com/share/5113opjozh).^101^ Despite being the largest to date (with a SNP-based heritability of 0.139 based on linkage-disequilibrium score regression analysis),^102^ the PCG GWAS only yielded two loci associated at genome-wide significance (p<5×10^-8^), near the *UGT1A8* (UDP Glucuronosyltransferase Family 1 Member A8) and *XPO6* (Exportin 6) genes (https://www.ebi.ac.uk/gwas/studies/GCST90103136). To have sufficient genetic instruments for causal inference, we applied a less stringent suggestive GWAS p-value threshold (p<5×10^-6^) to define independent instrumental variables not in linkage disequilibrium (LD r^2^ threshold = 0.05). Of note, the GSMR2 method accounts for the remaining LD not removed by LD clumping analysis.^32^ We used individual-level genotyping data from European (CEU) participants of the 1000 Genomes Project (Phase 3) as a reference to estimate LD structure.^103^

MR sensitivity analysis was performed to assess the potential confounding effect of kidney function on the association between PCG and cardiovascular outcomes. Using multi-trait-based conditional and joint analysis (mtCOJO, https://yanglab.westlake.edu.cn/software/gcta/#mtCOJO),^32^ we adjusted the PCG GWAS for genetic effects associated with estimated glomerular filtration rates (eGFR), which was adopted as a proxy for kidney function. Conditioning on a variable removes the non-causal association and controls for confounding. This conditional analysis required only GWAS summary data on the risk factor (i.e., eGFR), derived from an independent cohort of 1,004,040 individuals of European genetic ancestry background. Weights for eGFR genetic effects were also obtained from the GWAS Catalog (Supplementary Table 19). The conditional PCG GWAS summary statistics were then used for the MR sensitivity analysis.

### HAMSAB Cohort

Human ethics approval was obtained from the Monash University Human Research Ethics Committee (Study ID: 19203) and all participants signed informed consent. The study was registered with the Australian New Zealand Clinical Trials Registry (ACTRN12619000916145) and conducted following the Declaration of Helsinki. A detailed protocol on the rationale and study design was previously published.^34,104^ Briefly, this study was a double-blind, randomised, placebo-controlled cross-over trial conducted in Melbourne, Australia, at Monash University.

#### a) Inclusion criteria

Individuals with untreated hypertension were recruited by the study coordinator and screened for eligibility before being randomised. Inclusion criteria included being untreated for hypertension (as defined by the Australian National Heart Foundation guidelines, determined during the first study visit with 24-hour systolic BP[≥[130[mmHg and/or 24-hour diastolic BP[≥[80[mmHg), being of either sex (self-reported), 18–70 years of age and having a BMI of 18.5–35 kg/m^2^.

#### b) Exclusion criteria

Exclusion criteria included taking any anti-hypertensive medication, an office BP measurement >165/100[mmHg, antibiotic treatment in the last 3 months or probiotic intake over the previous 6 weeks. Those presenting with comorbidities (including type 1 or type 2 diabetes or any gastrointestinal disease), pregnancy or specific dietary requirements (for example, plant-based, gluten-free) or food intolerances were also excluded.

#### c) HAMSAB

This study utilised a modified version of high-amylose maize-resistant starch (HAMS), a type 2 resistant starch, which is considered highly fermentable by the gut microbiota, leading to the release of SCFAs.^105^ This supplement underwent further chemical modification by esterifying the SCFAs acetate and butyrate (HAMSAB). This modification ensured the targeted delivery and release of high levels of acetate and butyrate in the large intestines and systemic circulation^106,107^ and further fermentation of the residual HAMS in the colon.^107^

#### d) Intervention

Detailed intervention is described elsewhere.^34^ In brief, participants were assigned to either Diet A or Diet B for 3 weeks after successful screening and randomisation. Diet A contained HAMSAB (obtained from Ingredion) and was delivered at 40g per day, divided into two portions: 20g in the morning meal and 20g in the evening meal. Diet B contained 40g per day of placebo (corn starch or regular flour with no added resistant starches), delivered similarly, for 3 weeks. All other nutritional components (including energy, fat, protein, and sodium) were the same. After a 3-week washout period, participants were switched to the opposite arm for 3 weeks. All study food was stored in a –20°C freezer at Monash University. Participants were given a list of foods that were naturally high in RS and SCFAs and instructed to avoid them for the duration of the study.

#### e) Blood sample collection and targeted metabolomics

Fasting blood was collected in the morning, at least 12 hours after the last HAMSAB or placebo meal. Plasma *p*-Cresol, PCG and PCS were quantified using a targeted approach by a Triple quadrupole 7500 mass spectrometer (AB Sciex, Foster City, CA, USA) coupled with a Nexera LC-30AD UHPLC (Shimadzu Corporation, Kyoto, Japan) system, in collaboration with Professor John O’Sullivan at the University of Sydney. The targeted metabolite profiling used in this study was established using reference standards for each metabolite to determine MS multiple reaction-monitoring (MRM) transitions as described previously.^108^ PCG (TRC C782005), PCS (Cambridge Isotope Laboratories, DLM-9786) and *p*-Cresol (Sigma-Aldrich C85751) chemical standards were used to optimise individual MRM transitions, decluttering potentials, collision energies and specific chromatographic retention times using a Waters Amide column (Acquity UPLC BEH Amide, 1.7um,2.1 x 100mm, Waters Corp., Milford, MA, USA). In brief, the chromatographic separation was achieved on an Amide column using a gradient program with mobile A containing 5% acetonitrile, 95% water, and 10 mM ammonium acetate with pH adjusted to 9; and mobile B containing 100 % acetonitrile. The gradient started at 85% B and decreased to 35% at 8 min, and then continued to decrease to 2% in the next minute at 9 min. It was maintained at 2% B for another two minutes before ramping back to 85% within one minute. The system was re-equilibrated at 85% for at least 3 more minutes before the next injection. The flow rate was operated between 0.2–0.5 mL/min, depending on elution requirements.

Sciex OS 3.1 (AB Sciex) analysis software was used for MRM Q1/Q3 peak integration of the raw data files. The peak area corresponds to the abundance of that metabolite, and the abundance values were then normalised to the pooled plasma extracts in the subsequent analysis to account for any temporal RT drift and/or MS variations in instrument performance. The statistical analysis was performed with a moderated t-test (LIMMA) adapted for the analysis of cross-over trial data.

## Reporting summary

Further information on research design is available in the Nature Portfolio Reporting Summary linked to this article.

## Data Availability

***Animal cohorts, 16s and shotgun metagenome*:** Data will be deposited in a public database.

***VicGut metagenome data:*** Data will be deposited in a public database.

***TwinsUK cohort:*** The TwinsUK data used in this study are held by the Department of Twin Research at King’s College London. The data can be released to bona fide researchers using our normal procedures overseen by the Wellcome Trust and its guidelines as part of our core funding (http://twinsuk.ac.uk/resources-for-researchers/access-our-data/). MultiMuTHER RNASeq data are deposited in the European Genome-phenome Archive (EGA) (accession number EGA50000000135).

## Code Availability

The codes supporting this study’s findings are available from the corresponding author. Please email F.Z.M. at

## Acknowledgements

We acknowledge the Monash Proteomics and Metabolomics Platform for providing metabolomic profiling and measurement services, supported by BPA-enabled infrastructure through Bioplatforms Australia and the National Collaborative Research Infrastructure Strategy (NCRIS). We also thank Monash eResearch for access to the M3 servers, Research Data Storage, and the Nectar Research Cloud. We acknowledge Dr. Matthew Snelson and Dr. Tenghao Zheng for their guidance on microbiome analyses. We acknowledge and thank Dr. Chad Johnson for developing the macros used to analyse DAB (3,3’-diaminobenzidine) immunohistochemistry images, and more broadly, the Monash University Animal Research Platform, Monash University Histology Platform, Monash Bioinformatics Platform, and the Australian Genome Research Facility. For the VicGut study, we are grateful to Donna Vizi, Vivian Mak, and Kaye Carter, who assisted with blood collection. We also acknowledge Professor Graham Giles of the Cancer Epidemiology & Intelligence Division, Cancer Council Victoria, for permission to use the Dietary Questionnaire for Epidemiological Studies (Version 3.2), Melbourne: Cancer Council Victoria, 1996.

## Author information

**Authors and Affiliations**

**Hypertension Research Laboratory, Department of Pharmacology, Biomedical Discovery Institute, Faculty of Medicine, Nursing and Health Sciences, Monash University, Melbourne, Australia**

Chudan Xu, Liang Xie, Leticia Camargo Tavares, Evany Dinakis, Chaoran Yang, Michael Nakai, Joanne A. O’Donnell, Francine Z. Marques

**Victorian Heart Institute, Monash University, Melbourne, Australia**

Chudan Xu, Liang Xie, Leticia Camargo Tavares, Evany Dinakis, Chaoran Yang, Joanne A. O’Donnell, Francine Z. Marques

**Department of Obstetrics & Gynaecology, Yong Loo Lin School of Medicine, National University of Singapore, Singapore**

Liang Xie

**Monash Proteomics and Metabolomics Platform, Monash University, Melbourne, Australia**

Christopher K. Barlow

**Department of Biochemistry, Monash Biomedicine Discovery Institute, Monash University, Melbourne, Australia**

Christopher K. Barlow

**Department of Twin Research, King’s College London, UK**

Panayiotis Louca, Julia El-Sayed Moustafa, Kerrin Small, Cristina Menni

**CardioMetabolomic Medicine Group, School of Medical Sciences, Faculty of Medicine and Health, Charles Perkins Centre, The University of Sydney, Australia**

Xiaosuo Wang, Giovanni Guglielmi, John O’Sullivan

**Department of Gastroenterology, School of Translational Medicine, Monash University, Melbourne, Australia**

Dakota Rhys-Jones, Jane Muir

**Baker Heart and Diabetes Institute, Melbourne, Australia**

Stephanie Yiallourou, Melinda J. Carrington, David M. Kaye, Francine Z. Marques

**Iverson Health Innovation Research Institute and School of Health Sciences, Swinburne University of Technology, Hawthorn, Australia**

Gavin W. Lambert

**School of Pharmaceutical Sciences, Shandong Analysis and Test Center, Qilu University of Technology (Shandong Academy of Sciences), Jinan, China**

Charles R. Mackay

**Drug Delivery, Disposition & Dynamics, Monash Institute of Pharmaceutical Sciences, Monash University, Melbourne, Australia**

Darren J. Creek

**Department of Cardiology, Alfred Hospital, Melbourne, Australia**

David M. Kaye

**Monash-Alfred-Baker Centre for Cardiovascular Research, Monash University, Melbourne, Australia**

David M. Kaye

**Department of Cardiology, Royal Prince Alfred Hospital, Sydney, Australia**

John O’Sullivan

**Department of Pathophysiology and Transplantation, Università Degli Studi di Milano, Italy**

Cristina Menni

**Fondazione IRCCS Cà Granda Ospedale Maggiore Policlinico, Angelo Bianchi Bonomi Hemophilia and Thrombosis Center, Italy**

Cristina Menni

## Funding Information

This work was supported by a Senior Medical Research Fellowship from the Sylvia and Charles Viertel Charitable Foundation, and National Health & Medical Research Council (NHMRC) Emerging Leader Fellowship (GNT2017382) to F.Z.M. C.X. was supported by the Monash Graduate Scholarship and Postgraduate Publication Award from Monash University. L.X. is supported by a Young Individual Research Grant from National Medical Research Council of Singapore (MOH-001714). P.L. is funded by the Chronic Disease Research Foundation. J.O.D. is supported by an NHMRC Fellowship (GNT1124288). G.W.L. is supported by discretionary funding from Swinburne Research, Swinburne University of Technology. C.M. is funded by the Chronic Disease Research Foundation, by the UKRI (MR/Y010175/1, MR/T004142/1), the Italian Ministry of Education and Research: Dipartimenti di Eccellenza Program 2023 to 2027 and by the Italian Ministry of Health – Bando Ricerca Corrente. The Department of Twin Research received support from grants from the Wellcome Trust (212904/Z/18/Z) and the Medical Research Council (MRC) / British Heart Foundation (BHF) Ancestry and Biological Informative Markers for Stratification of Hypertension (AIM-HY; MR/M016560/1), European Union, Chronic Disease Research Foundation (CDRF), Zoe Global Ltd., the NIHR Clinical Research Facility and Biomedical Research Centre (based at Guy’s and St Thomas’ NHS Foundation Trust in partnership with King’s College London). The TwinsUK study was also supported by grant funding from Tinnitus UK. F.Z.M. is supported by a National Heart Foundation Future Leader Fellowship (105663). The Baker Heart & Diabetes Institute is supported, in part, by the Victorian Government’s Operational Infrastructure Support Program. The HAMSAB RCT was supported by a National Heart Foundation Vanguard grant (102182), a National Health & Medical Research Council (NHMRC) of Australia project grant (GNT1159721).

## Author Contributions

C.X. contributed to the conceptualisation, performed most of the experiments, collected data, carried out data analyses, and wrote the original manuscript, with input from F.Z.M. L.X. contributed to the animal studies and data collection. C.K.B. performed the metabolomic profiling and the processing of raw data. L.C.T. contributed to Mendelian Randomisation and shotgun metagenomic analyses and data presentation. E.D. helped with animal studies and data collection. P.L., J.E.M., K.S. and C.M. contributed to the TwinsUK studies, resource availability and data analyses. C.Y. contributed to RNA-sequencing analyses and output visualisation. M.N. contributed to gut microbiome library preparation and the processing of raw data. D.R.J. coordinated the HAMSAB trial, recruited participants and collected samples and data. J.M. contributed to the conception, design and supervision of the HAMSAB trial, and critically revised it and contributed to data interpretation. X.W., G.G. and J.O.S. performed the targeted metabolomics on the HAMSAB study and the downstream data analyses. J.O.D. contributed to conceptualisation. S.Y. and M.J.C. coordinated the VicGut study, recruited participants and collected samples and data. G.W.L. performed high-performance liquid chromatography to quantify catecholamines, processed the raw data, reviewed and approved the final manuscript. D.J.C. supervised the metabolomics analyses.

C.R.M. provided HAMSAB for the trial. D.M.K. contributed with mouse (low-vs high-fibre) and human samples (VicGut). F.Z.M. conceived and designed the study, supervised the project, secured funding, and contributed towards the writing. All authors had full access to all data in the study, revised the manuscript critically, approved the version to be published and had final responsibility for the decision to submit for publication.

## Conflict of interest

The authors declare no competing interests.

## Notes

### Competing Interest Statement

The authors have declared no competing interest.

### Clinical Trial

ACTRN12619000916145, ACTRN12620000958987

### Author Declarations

The study complied with the Declaration of Helsinki and was approved by the Alfred Hospital human research ethics committee (approval 415/16). Ethical approval for the TwinsUK study was granted by the St Thomas Hospital Research Ethics Committee, and all participants provided written informed consent (REC ref EC04/015). Human ethics approval was obtained from the Monash University Human Research Ethics Committee (Study ID: 19203) and all participants signed informed consent.

