## Supplementary material for "Lack of dietary fibre increases gut microbiome-derived uremic toxins that contribute to increased blood pressure": Online supplementary figures

**Supplementary Figure 1.** Dietary fibre modulated the mouse gut microbiome.

**Supplementary Figure 2.** The effect of dietary fibre on the mouse gut metagenome.

**Supplementary Figure 3.** Validation the identification of *p*-Cresol glucuronide and *p*-Cresol sulfate.

**Supplementary Figure 4.** The impact of a lack of dietary fibre and Ang II-induced hypertension on mouse metabolic pathways.

**Supplementary Figure 5.** L-Tyrosine supplementation did not change plasma DOPA *in vivo*.

**Supplementary Figure 6.** Mendelian Randomisation between PCG and blood pressure based on multi-trait based conditional and joint (mtCOJO) analysis.

**Supplementary Figure 7.** Genes positively correlated with *p*-Cresol-derived metabolites.

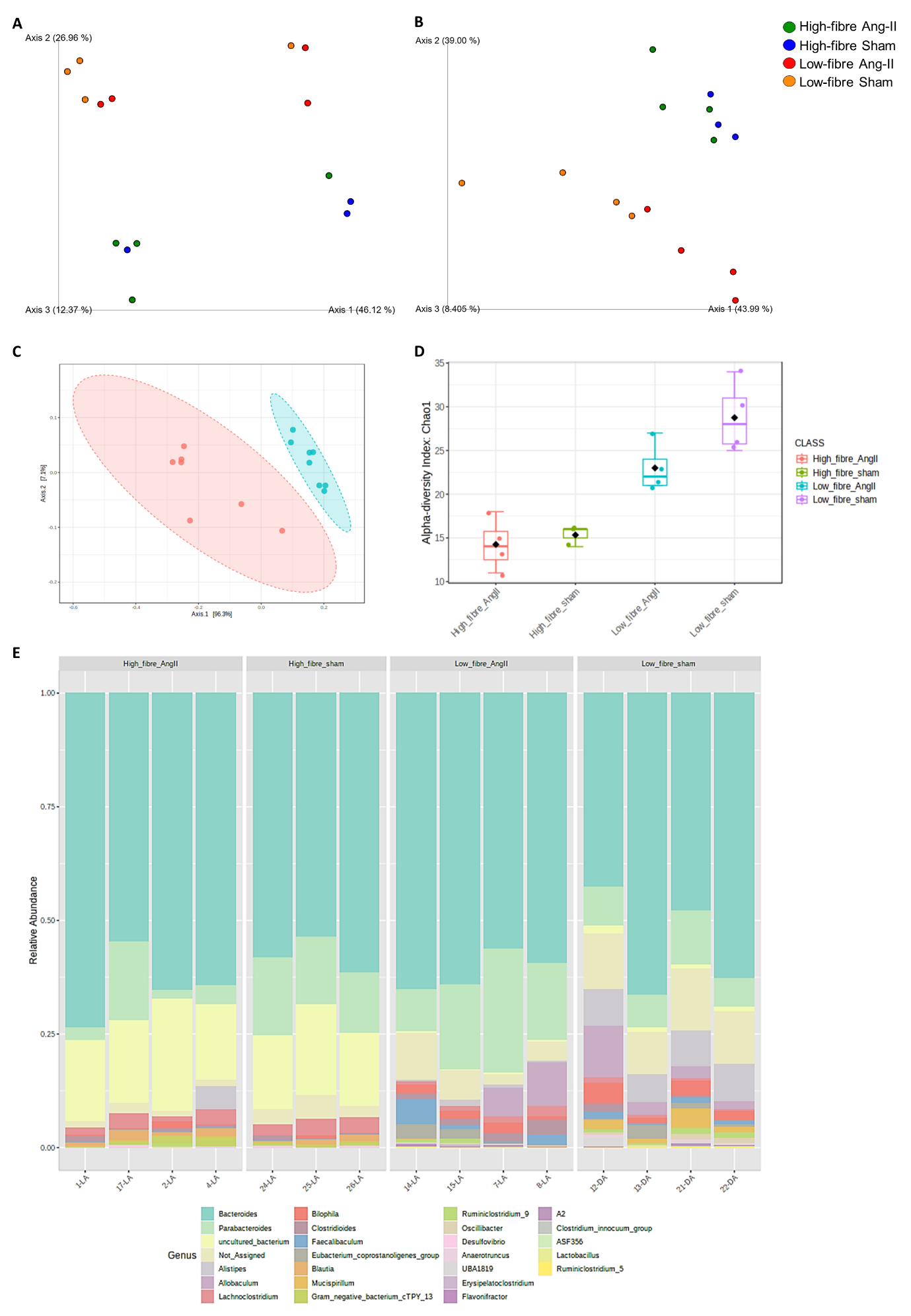

**Supplementary Figure 1. Dietary fibre modulated the mouse** **gut microbiome.** Caecal samples were extracted from Ang II-induced hypertensive mice and normotensive control mice fed either a low- or high-fibre diet. Caecal DNA were sequenced using 16S rRNA gene sequencing technique. Principal coordinates analyses with **(A)** unweighted and **(B)** weighted taxonomic metrics. **(C)** β-diversity between low- and high-fibre diet groups using Jensen-Shannon divergence distance method at the species level (n=7-8/group). Bacterial relative abundance at the **(D)** phylum and **(E)** genus level (n=3-4/group)**.** Gut microbiome analyses were conducted using QIIME2 in R Studio and MicrobiomeAnalyst (version 2.0).

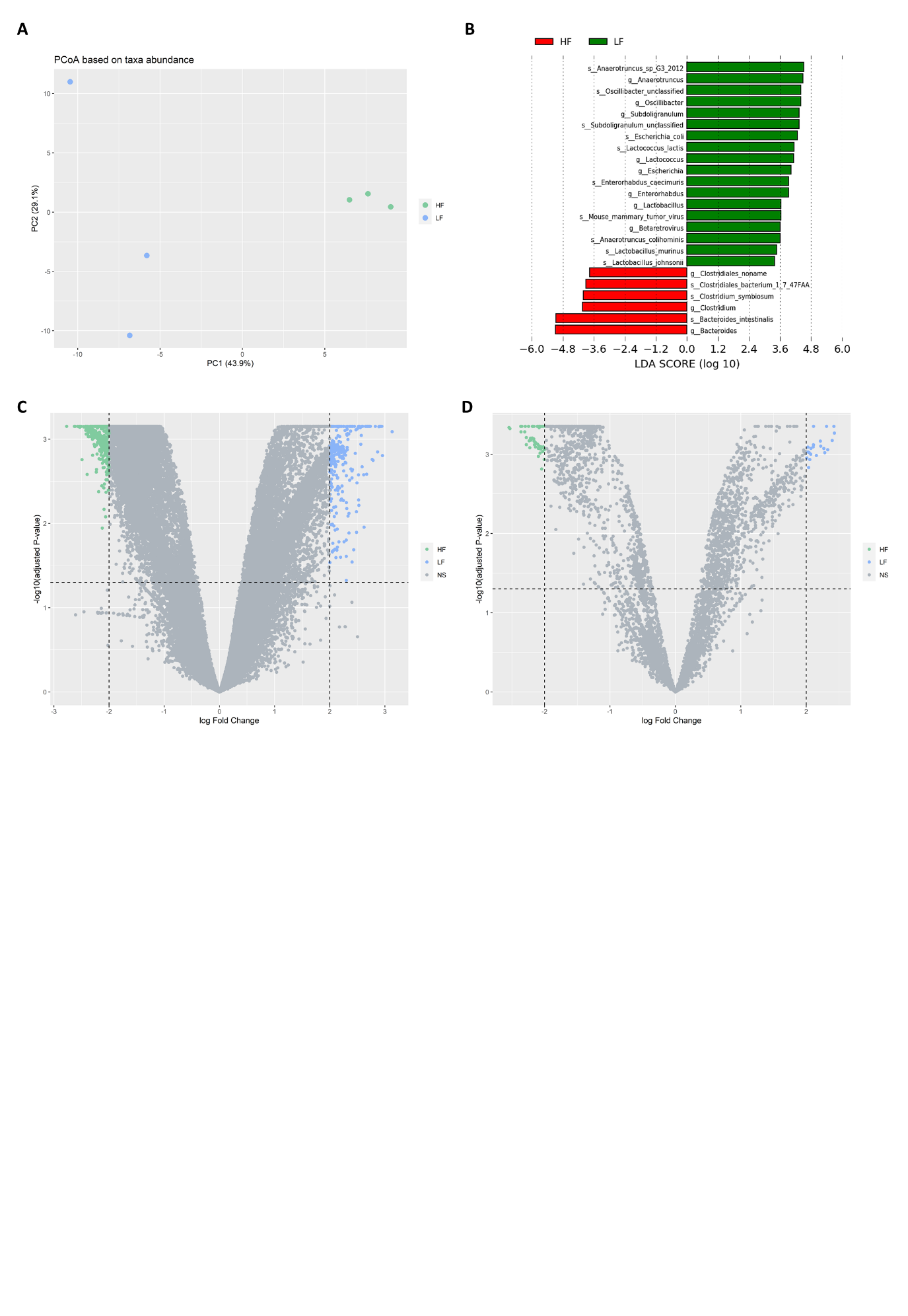

**Supplementary Figure 2. The effect of dietary fibre on the mouse gut metagenome.** Composition of the intestinal microbiota (caecum content) of low- and high-fibre intake mice from shotgun sequencing. An overall representation of microbial profiles in low-fibre (0%RS) and high-fibre (62%RS) mice at the endpoint (12-week age-old). **(A)** Principal coordinates analyses based on differential abundance of taxa. **(B)** Linear discriminant analysis (LDA) effect size (LEfSe) analyses at the species level. Volcano plot showing differential abundance in **(C)** gene family and **(D)** enzyme commission between low- and high-fibre intake. n=3 pool sample per group (1-2 biological independent samples pooled together). The differential abundance of taxa was analysed using the ALDEx2 package (version 1.22.0). LEfSe analyses were performed through the Galaxy platform (Afgan et al., 2018). Gene family (UniRef) and enzyme commission category-grouped genes (KEEG) abundance counts were analysed for differential expression between diets using the limma package (version 3.46.0). Linear discriminant analysis (LDA) effect size (LEfSe) analyses were performed in the Galaxy platform. The LDA scores of the high fibre were negative, while those of the low fibre were positive. Such negativity or positivity was determined by alphabetical order of the groups, and the absolute values of the effect size indicate the scale of the difference between the two groups, regardless of the positivity or negativity.

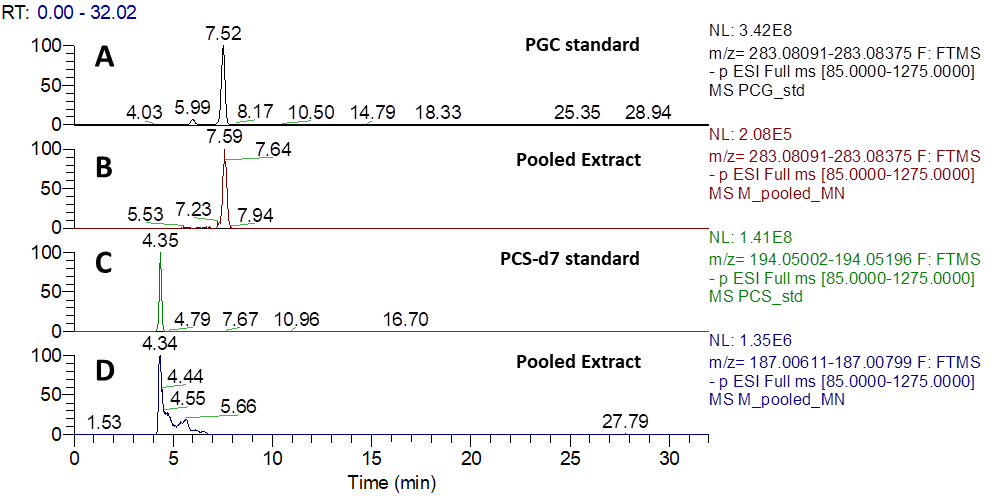

**Supplementary Figure 3.** **Validation of the identification of *p*-Cresol glucuronide and *p*-Cresol sulfate.** The extracted ion chromatograms (EIC) at the m/z corresponding to the PCG anion for a PCG standard (**A**) and a pooled mouse plasma extract (**B**). Here we observe good agreements in the retention time of the standard and the feature in the pooled extract around 7.5 to 7.6 minutes, supporting the assignment of PCG from the untargeted analysis. The EIC at the m/z for the deuterated PGS for a PCS-d7 standard (**C**), and the EIC at the m/z for the unlabelled PCS for a pooled mouse plasma extract (**D**). Similarly, we observe a feature in both cases around 4.3 minutes supporting the assignment of PCS.

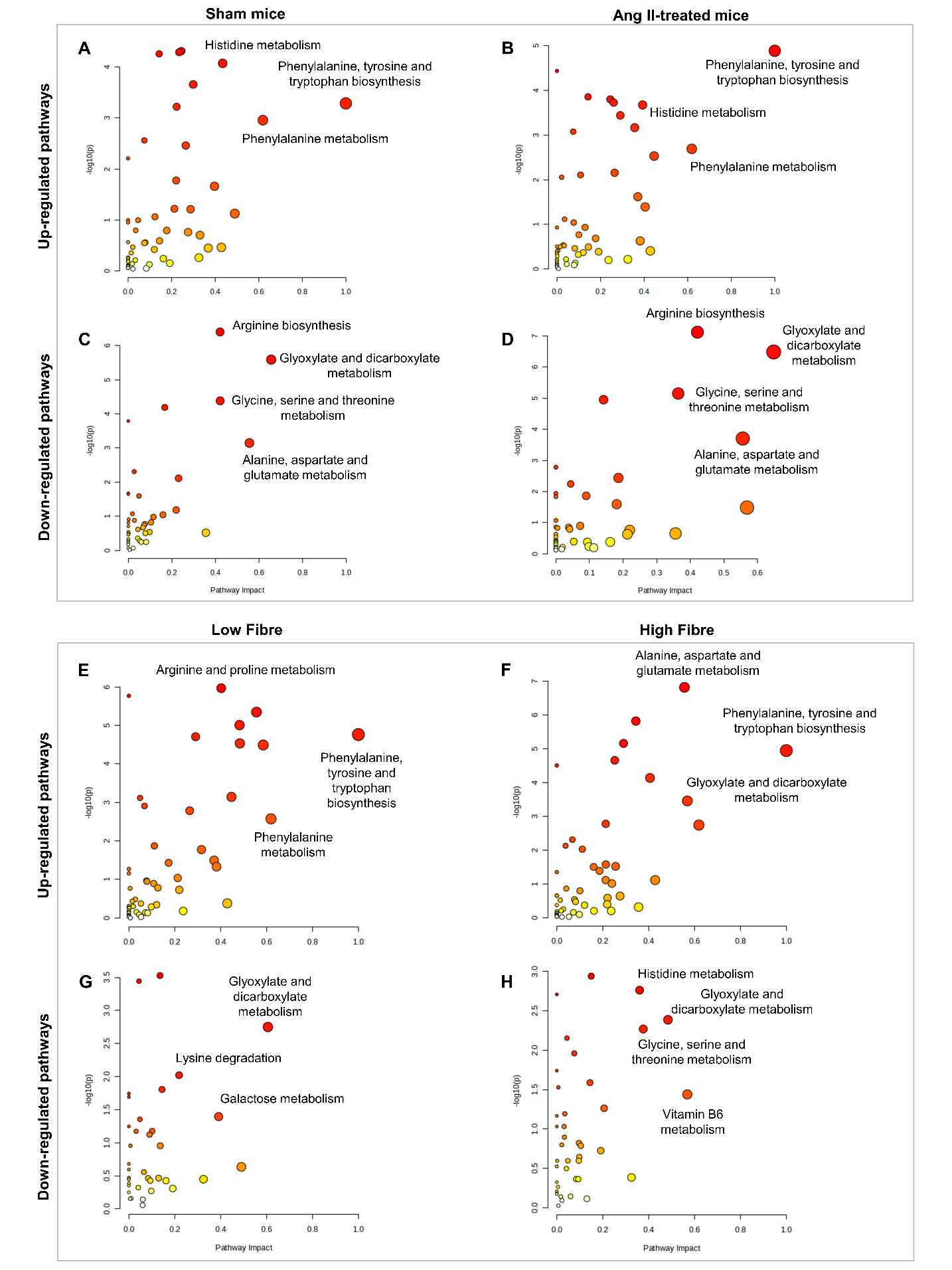

**Supplementary Figure 4. The impact of a lack of dietary fibre and Ang II-induced hypertension on mouse metabolic pathways.** Mice had either low- or high-fibre dietary intervention at three-week-old of age (WOA). At 6 WOA, they had minipump surgery and received either Angiotensin II intervention (hypertensive group) or saline (sham/normotensive group) (n=3-4/group). After 4 weeks of intervention, plasma was collected and used for metabolomic profiling. The impact of fibre deficient on metabolic pathways: up-regulated pathways in response to low- versus high-fibre intake in **(A)** sham and **(B)** Ang II-treated mice. Down-regulated pathways in response to low- versus high-fibre intake in **(C)** sham and **(D)** Ang II-treated mice. The effect of Ang II on the metabolic pathway: up-regulated pathways in Ang II-induced hypertensive mice compared to sham mice under **(E)** a low-fibre and **(F)** a high-fibre diet. Down-regulated pathways in Ang II-induced hypertensive mice compared to sham mice under **(G)** a low-fibre and **(H)** a high-fibre diet. Analyses were performed via MetaboAnalyst using one-way ANOVA, FDR<0.05 and |log2FC|≥1. The pathway enrichment analysis was based on KEGG Pathway. Ang II, angiotensin II. KEGG, Kyoto Encyclopedia of Genes and Genomes.

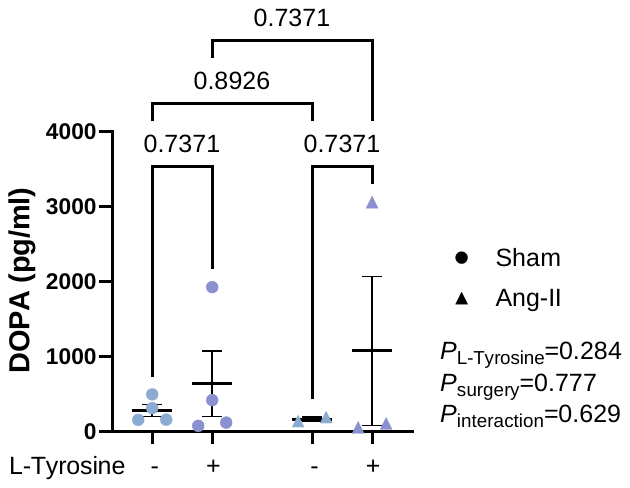

**Supplementary Figure 5. L-Tyrosine supplementation did not change plasma DOPA *in vivo*.** The level of DOPA (pg/ml) in the circulation. Two-way ANOVA adjusted for multiple comparisons (n=4-6/group). All values are expressed as mean ±SEM.

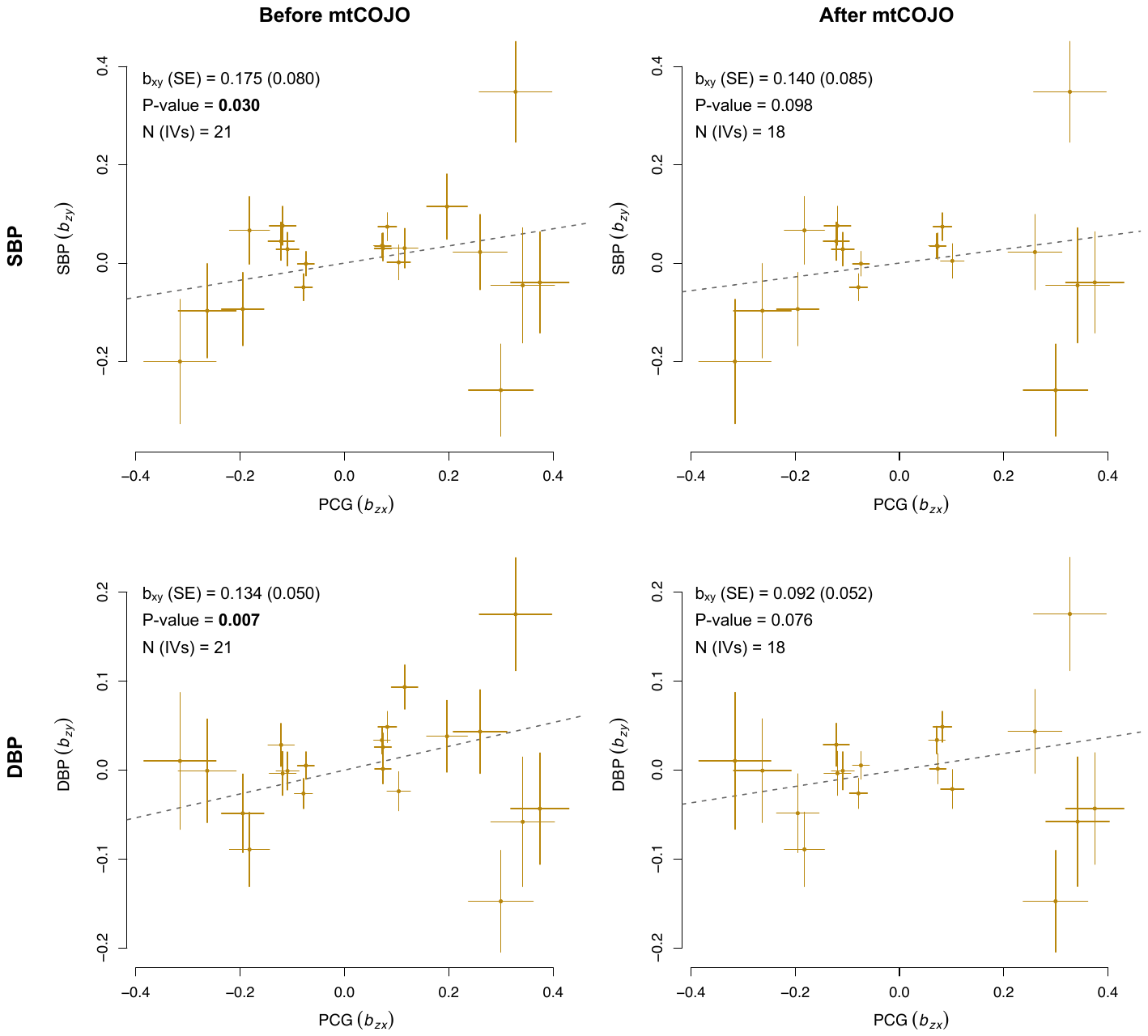

**Supplementary Figure 6. Mendelian Randomisation between PCG and blood pressure based on multi-trait based conditional and joint (mtCOJO) analysis.** MR results between PCG as exposure and SBP/DBP as outcomes, before and after conditioning on eGFR genetic effects based on mtCOJO analysis. The figure depicts the effects of each selected genetic instrumental variable (IVs) on the exposure (b_zx_) and outcomes (b_zy_), as well as the overall MR summary statistics results (b_xy_ and P-value). Full results in **Table 5**. PCG: *p*-cresol glucuronide; SBP: systolic blood pressure; DBP: diastolic blood pressure; eGFR: estimated glomerular filtration rate.

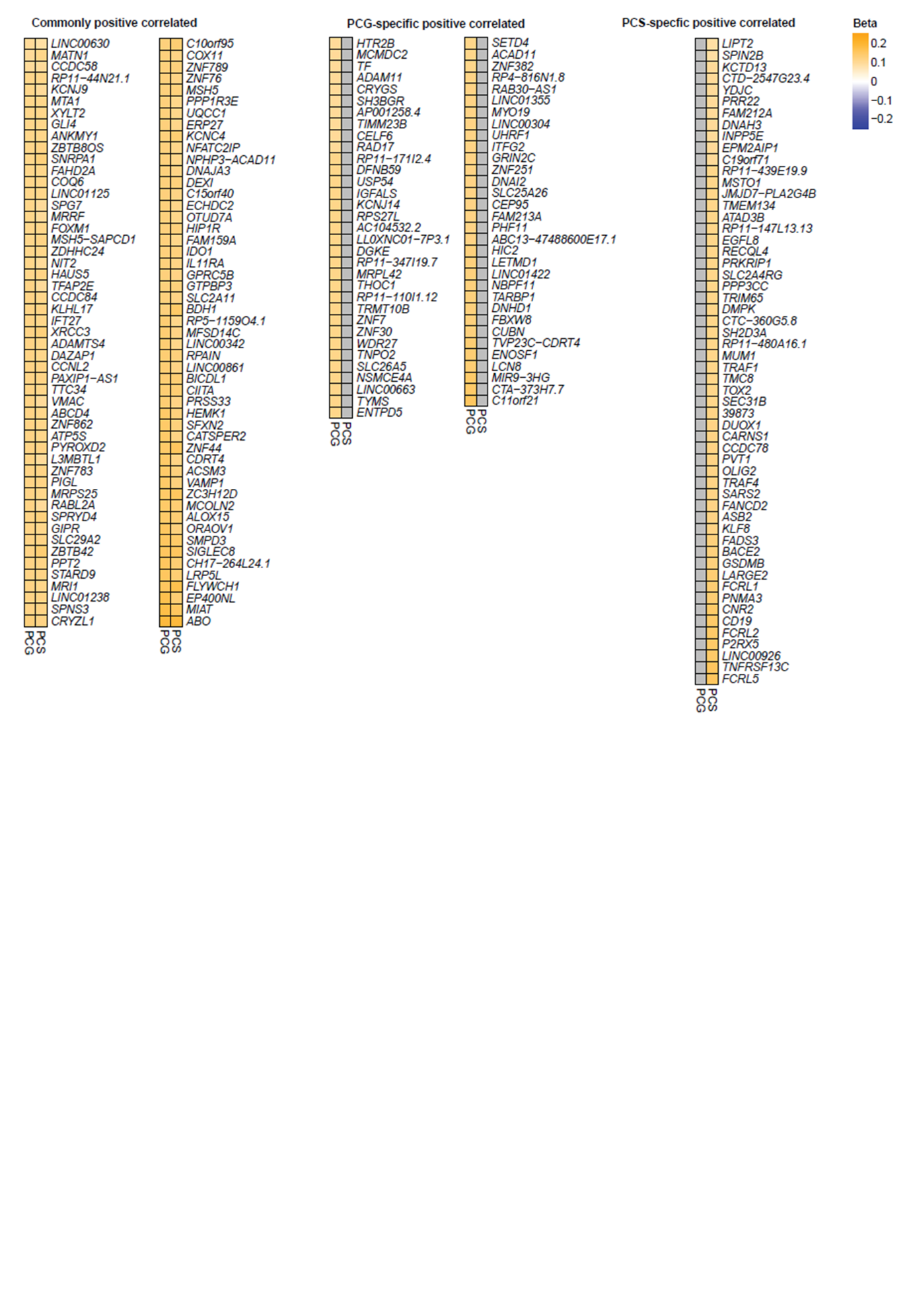

**Supplementary Figure 7.** **Genes positively correlated with *p*-Cresol-derived metabolites.** Co-expression analyses were performed between blood RNA-sequencing and *p*-Cresol-derived metabolites in the TwinsUK cohort (n=933). Genes positively correlated to both *p*-Cresol glucuronide (PCG) and *p*-Cresol sulfate (PCS).
